# Seroconversion stages COVID19 into distinct pathophysiological states

**DOI:** 10.1101/2020.12.05.20244442

**Authors:** Matthew D. Galbraith, Kohl T. Kinning, Kelly D. Sullivan, Ryan Baxter, Paula Araya, Kimberly R. Jordan, Seth Russell, Keith P. Smith, Ross E. Granrath, Jessica Shaw, Monika Dzieciatkowska, Tusharkanti Ghosh, Andrew A. Monte, Angelo D’Alessandro, Kirk C. Hansen, Tellen D. Bennett, Elena W.Y. Hsieh, Joaquin M. Espinosa

## Abstract

COVID19 is a heterogeneous medical condition involving a suite of underlying pathophysiological processes including hyperinflammation, endothelial damage, thrombotic microangiopathy, and end-organ damage. Limited knowledge about the molecular mechanisms driving these processes and lack of staging biomarkers hamper the ability to stratify patients for targeted therapeutics. We report here the results of a cross-sectional multi-omics analysis of hospitalized COVID19 patients revealing that seroconversion status associates with distinct underlying pathophysiological states. Seronegative COVID19 patients harbor hyperactive T cells and NK cells, high levels of IFN alpha, gamma and lambda ligands, markers of systemic complement activation, neutropenia, lymphopenia and thrombocytopenia. In seropositive patients, all of these processes are attenuated, observing instead increases in B cell subsets, emergency hematopoiesis, increased markers of platelet activation, and hypoalbuminemia. We propose that seroconversion status could potentially be used as a biosignature to stratify patients for therapeutic intervention and to inform analysis of clinical trial results in heterogenous patient populations.

## INTRODUCTION

COVID19 (coronavirus disease of 2019), the disease caused by the severe acute respiratory syndrome coronavirus 2 (SARS-CoV-2), has caused more than 1.48 million deaths worldwide since late 2019. SARS-CoV-2 is a highly contagious coronavirus that uses angiotensin-converting enzyme-2 (ACE-2), a protein widely expressed on lung type II alveolar cells, endothelial cells, enterocytes, and arterial smooth muscle cells, as its primary cellular entry receptor (1). Neuropilin-1 (NRP1) has been characterized as an additional entry receptor for SARS-CoV-2, thus extending the range of host cells and tissues directly affected by the virus (2, 3). The clinical presentation of COVID19 is highly variable, ranging from asymptomatic infection to multiorgan failure and death (4). Mild symptoms include a flu-like condition consisting of fever, nasal congestion, cough, fatigue, and myalgia. In a small fraction of patients, SARS-CoV-2 causes more severe effects in multiple organ systems. These include respiratory failure, thromboembolic disease, thrombotic microangiopathies, stroke, neurological symptoms including seizures, as well as kidney and myocardial damage (4). The molecular and cellular bases of this clinical heterogeneity remain to be elucidated.

Several pathophysiological processes have been implicated in the etiology of severe COVID19 symptoms, including but not restricted to a hyperinflammation-driven pathology (5), disruption of lung barrier function by Type I and III interferons (IFN) (6, 7), organ damage by systemic activation of the complement cascade (8), vascular pathology caused by a bradykinin storm (9), and a dysregulated fibrinolytic system (10). The interplay between these non-mutually exclusive processes is yet to be fully elucidated, and each of them offers opportunities for therapeutic interventions currently being tested in clinical trials. However, the lack of precise biomarkers for cohort stratification and targeted therapeutics has hampered progress in this area.

We report here the results of a cross-sectional multi-omics analysis of hospitalized COVID19 patients investigating the multidimensional impacts of seroconversion status. When stratifying patients by a quantitative metric of seroconversion, or ‘seroconversion index’, we were able to define biosignatures differentially associated with humoral immunity. Low seroconversion indices associate with high levels of activated T cells and cytokine-producing Natural Killer (NK) cells, biosignatures of monocyte activation, high levels of IFN alpha, gamma and lambda ligands, markers of systemic complement activation, neutropenia, lymphopenia, and thrombocytopenia. In seroconverted patients, all these biosignatures are decreased or fully reversed, leading instead to increased levels of circulating plasmablasts and mature and activated B cell subsets, increased numbers of neutrophils, lymphocytes and platelets, elevated markers of platelet degranulation, and significantly decreased levels albumin and major liver-derived proteins, indicative of vascular leakage. Altogether, these results indicate that a quantitative assessment of seroconversion status could be employed to map the trajectory of underlying pathophysiological processes, with potential utility in stratification of patients in the clinic and enhanced interpretation of clinical trial data.

## RESULTS

### Hospitalized COVID19 patients display highly variable seroconversion status

In order to investigate variations in the pathophysiological state of COVID19 patients, we completed an integrated analysis of 105 research participants, including 73 COVID19 patients versus 32 SARS-CoV-2 negative controls (**Fig. 1a**). Cohort characteristics are summarized in **Supp. File S1**. COVID19 patients tested positive for SARS-CoV-2 infection by PCR and/or antibody testing and were hospitalized due to COVID19 symptoms, but none of them had developed severe pathology requiring ICU admission at the time of blood collection. The control group was recruited from the same hospital system but tested negative for SARS-CoV-2 infection. Research blood draws were obtained from consented participants and analyzed by matched SARS-CoV-2 seroconversion assays, plasma proteomics using two alternative platforms [mass-spectrometry (MS) and SOMAscan® assays], 82-plex cytokine profiling using multiplex immunoassays with Meso Scale Discovery (MSD) technology, and immune cell profiling via mass cytometry (MC) (**Fig. 1a**) (see Methods).

**Figure 1.**
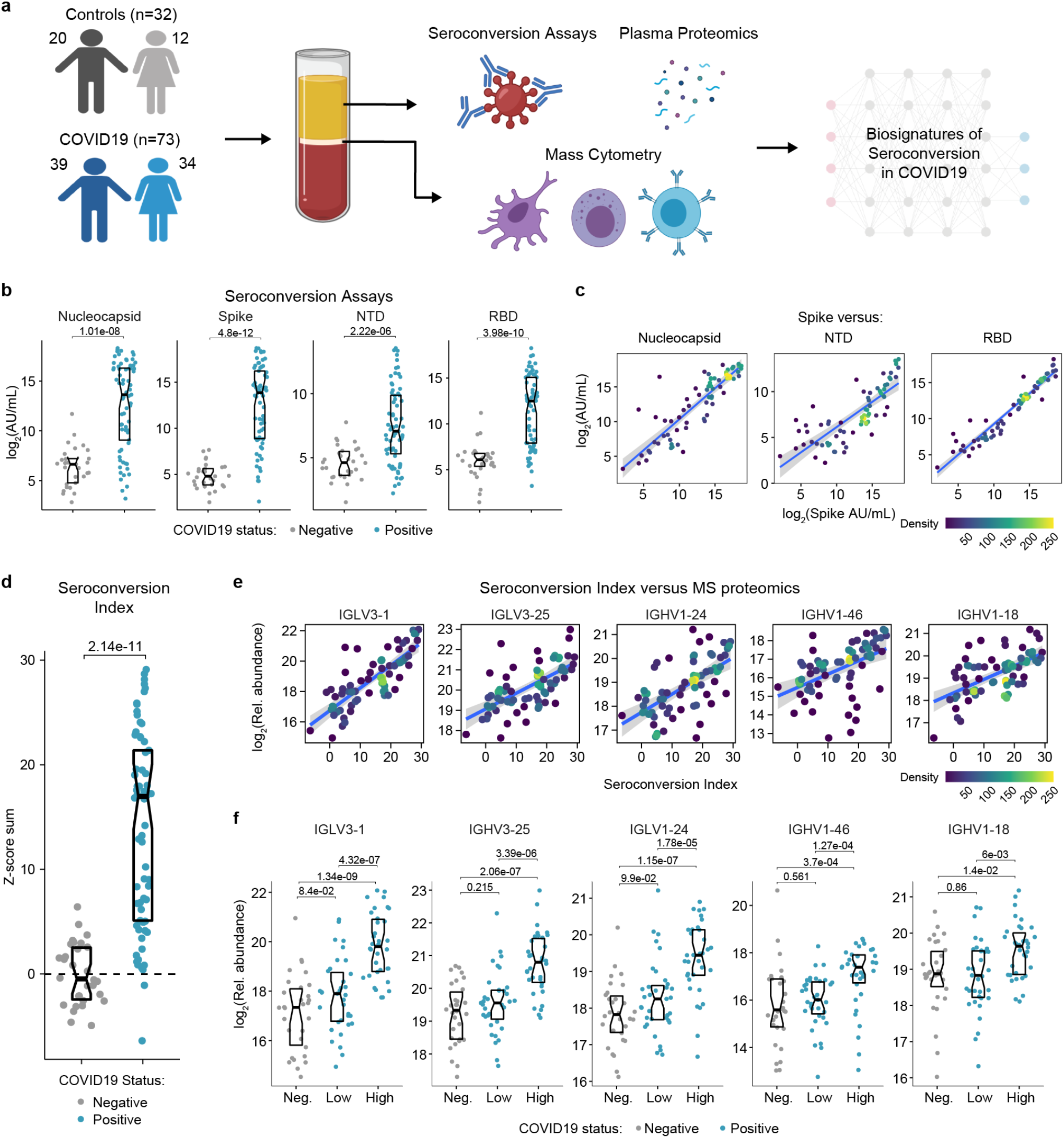
Highly variable seroconversion status among hospitalized COVID19 patients. **a**. Overview of experimental approach. Blood samples from 105 research participants, 73 of them with COVID19, were analyzed by matched multiplex immunoassays for detection of antibodies against SARS-CoV-2, plasma proteomics using mass spectrometry (MS), SOMAscan^®^ proteomics, and cytokine profiling using Meso Scale Discovery (MSD) technology. Data was then analyzed to define biosignatures of seroconversion. **b**. Multiplex immunoassays were used to measure antibodies against the SARS-CoV-2 nucleocapsid and spike proteins, as well as specific peptides encompassing the N-terminus domain (NTD) and receptor-binding domain (RBD) of the spike protein. Data are presented as modified Sina plots with boxes indicating median and interquartile range. Numbers above brackets are p-values for Mann-Whitney tests. **c**. Scatter plots showing correlations between antibodies against the full-length spike protein versus antibodies against the nucleocapsid, NTD and RBD domains. Points are colored by density; lines represent linear model fit with 95% confidence interval. **d**. Seroconversion indices were calculated for each research participant by summing the Z-scores for each of the four seroconversion assays. **e**. Scatter plots displaying the top five correlations between seroconversion indices and proteins detected in the MS proteomics dataset among COVID19 patients. Points are colored by density; lines represent linear model fit with 95% confidence interval. **f**. Sina plots showing values for the top 5 proteins correlated with seroconversion comparing the control cohort (negative) to COVID19 patients divided into seroconversion low and high status. Data are presented as modified Sina plots with boxes indicating median and interquartile range. Numbers above brackets are Q-values for Mann-Whitney tests. See also **Figure S1**.

In order to stratify the COVID19 positive cohort, we measured seroconversion with multiplex immunoassays detecting IgGs against four different SARS-CoV-2 peptides: full length nucleocapsid, full length spike protein (spike), as well as smaller peptides encompassing the N-terminus domain (NTD) and the receptor binding domain (RBD) of the spike protein (see Methods). The COVID19 cohort displayed significantly elevated levels of anti-SARS-CoV-2 IgGs in all four assays, with strong inter-individual variability (**Fig. 1b**).

As a control, levels of antibodies against the Flu A Hong Kong H3 virus strain were no higher in COVID19 patients (**Fig. S1a**). In COVID19 patients, reactivity against the spike protein correlated positively with reactivity against the other three peptides (**Fig. 1c**). Therefore, we generated a seroconversion index by summing Z-scores for each individual seroconversion assay, which enabled us to assign a quantitative seroconversion value to each patient (**Fig. 1d**). We then set out to define biosignatures significantly associated, either positively or negatively, with the seroconversion index among COVID19 patients by analyzing correlations with the proteome, cytokine profiling, and MC datasets. When calculating Spearman correlation values between seroconversion indices and individual features in the other datasets, we identified hundreds of proteins and dozens of immune cell types significantly correlated with seroconversion (**Fig. S1b-f, Supp. Files S2-5**).

Reassuringly, top positive correlations among 407 abundant plasma proteins detected by MS are dominated by specific immunoglobulin sequences, including several that were previously observed to be enriched in the bloodstream of COVID19 patients during seroconversion (**Supp. File S2, Fig. 1e-f**) (11).

Altogether, these observations suggest that seroconversion is accompanied by significant changes in underlying pathophysiological processes in COVID19, which prompted us to complete a more thorough analysis of these correlations.

### Immune cell signatures of seroconversion in COVID19

First, we investigated associations between seroconversion and changes in the peripheral immune cell compartment among COVID19 patients. Among all live CD45+ white blood cells (WBCs), significant negative associations included plasmacytoid dendritic cells (pDCs), distinct subsets of CD4+ and CD8+ T cells, and CD56^bright^ NK cells (**Fig. 2a-b, Fig. S2a-b**). Conversely, positive associations were dominated by B cell subsets. pDCs were only mildly elevated in seronegative COVID19 patients relative to the control group but significantly decreased in the circulation of seropositive patients (**Fig. 2c**). Being first responders during a viral infection, pDCs are predicted to be activated and extravasate into the circulation early on as part of the innate immune response, ahead of development of humoral immunity. Their significant reduction in the bloodstream of seropositive patients could be indicative of exhaustion and/or depletion over the course of the disease. Among CD4+ T cells, we observed elevated circulating levels of Th1, Th17, Th1/17, follicular helper CD4+ T cells (T_FH_), and terminally differentiated effector memory CD45RA+ subsets in seronegative COVID19 patients, with levels falling back to baseline or below baseline in seropositive patients (**Fig. 2a,c**). Among CD8+ T cells, a similar behavior was observed for activated (CD95+), effector (T-bet+Eomes+), senescent (T-bet+ Eomes-), effector memory, and terminally differentiated CD45RA+ subsets (**Fig. 2a-c, Fig. S2a-b**). These patterns were largely conserved when calculating frequencies within all T cells and within CD4+ and CD8+ T cell subsets (**Fig. S2a, Supp. File S5**). These changes in peripheral T cell subsets are consistent with an acute and transient antiviral T cell response in patients with low seroconversion indices, marked by elevated levels of activated and effector CD8+ T cells, polarization of CD4+ T cells toward the Th1 antiviral state, accompanied by development of T cell memory, T_FH_-assisted maturation of B cells, and eventual senescence and terminal differentiation of cytotoxic CD8+ T cells. This bimodal T cell behavior is accompanied by increases in CD56^bright^ NK cells only in seronegative patients (**Fig. 2a,c**). CD56^bright^ NK cells lack expression of inhibitory receptors and express high levels of activating receptors, cytokine and chemokine receptors, and adhesion molecules (12). Although CD56^bright^ NK cells are not as cytotoxic as other NK subsets, they are strong producers of key cytokines involved in the immune response, most prominently IFNG, which we found to be elevated in seronegative patients (see later, **Fig. 3**). CD56^bright^ NK cells have been found to be elevated in some autoimmune conditions and infections (12).

**Figure 2.**
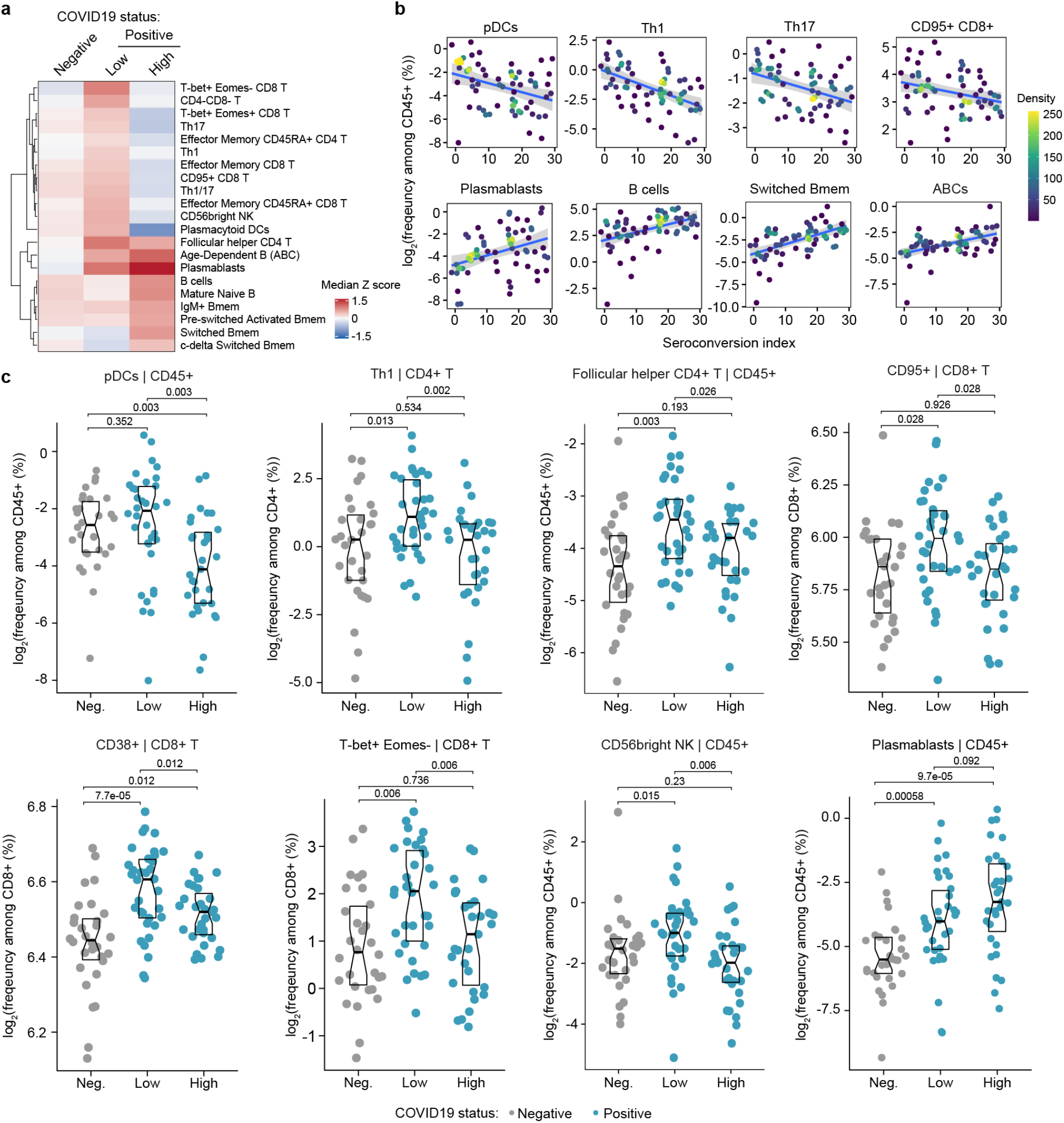
Seroconversion associates with significant changes in peripheral immune cell frequencies. **a**. Heatmap representing changes in immune cell subsets that are significantly correlated, either positively or negatively with seroconversion status. Values displayed are median Z-scores, derived from cell frequencies among all CD45+ cells, for each cell subset for controls (negative, Neg.) versus COVID19 patients divided into seroconversion low (Low) and high (High) status. See also **Figure S2a. b**. Scatter plots for indicated immune cell types positively and significantly correlated with seroconversion indices among COVID19 patients. Points are colored by density; lines represent linear model fit with 95% confidence interval. See also **Figure S2b. c**. Sina plots showing values for indicated immune cell types significantly correlated with seroconversion indices among COVID19 patients. The parent cell lineage is indicated in the header and Y axis label for each plot. Data are presented as modified Sina plots with boxes indicating median and interquartile range. Numbers above brackets are Q-values for Mann-Whitney tests. See also **Figure S2c**.

**Figure 3.**
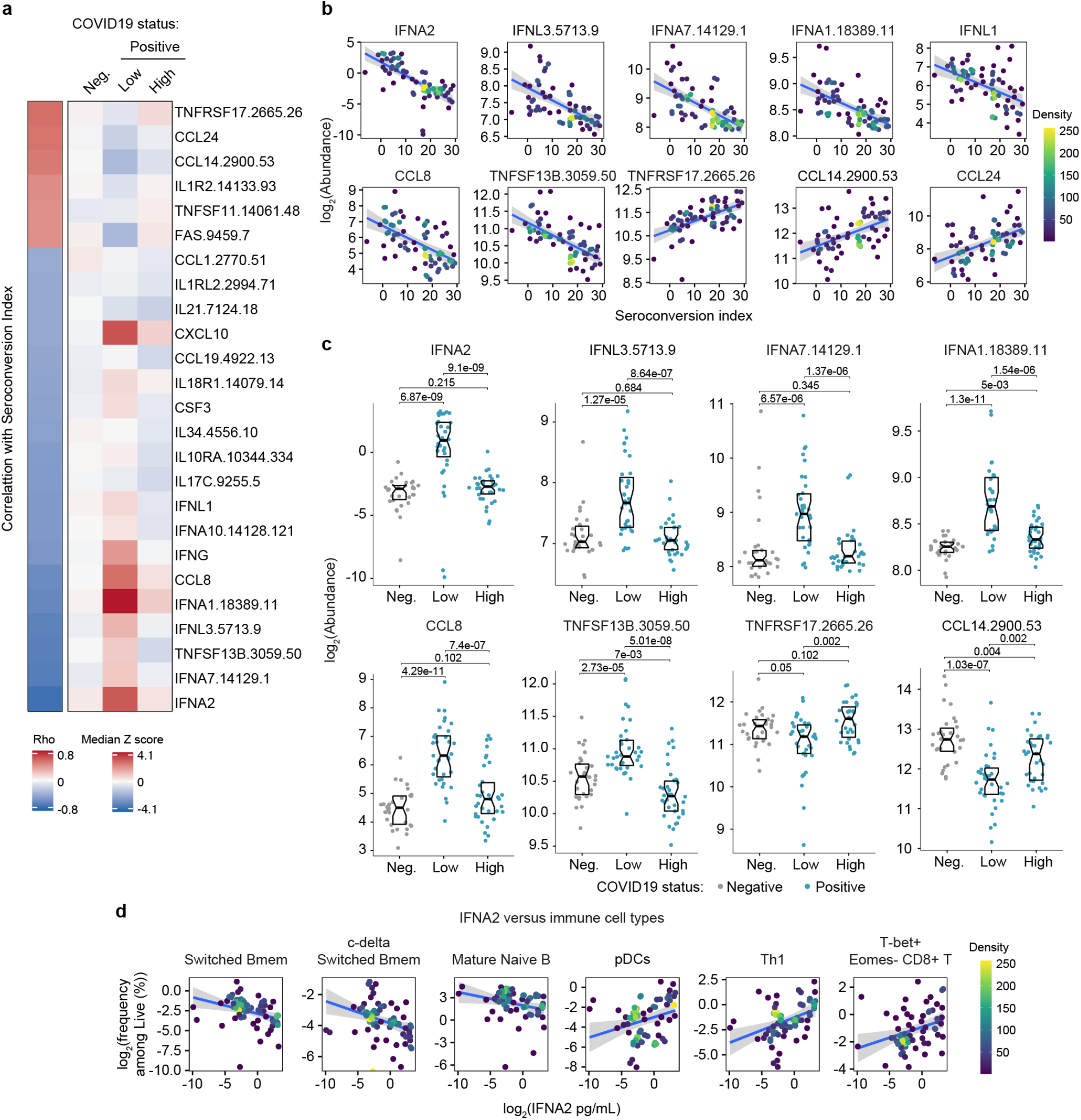
Seroconversion is associated with decreased interferon signaling. **a**. Heatmap displaying changes in circulating levels of immune factors that are significantly correlated, either positively or negatively, with seroconversion status. The left column represents Spearman *rho* values, while the right columns display median Z-scores for each immune factor for controls (negative, Neg.) versus COVID19 patients divided into seroconversion low (Low) and high (High) status. Factors are ranked from most positively correlated (top, high *rho* values) to most anti-correlated (bottom, low *rho* values) with seroconversion index. **b**. Scatter plots for indicated immune factors significantly correlated with seroconversion indices among COVID19 patients. Points are colored by density; lines represent linear model fit with 95% confidence interval. See also **Figure S3a. c**. Sina plots showing values for immune factors correlated with seroconversion comparing controls (Neg.) to COVID19 patients divided into seroconversion low and high status. Data are presented as modified Sina plots with boxes indicating median and interquartile range. Numbers above brackets are Q-values for Mann-Whitney tests. **d**. Scatter plots showing correlations between circulating levels of IFNA2 measured by MSD and the indicated cell types measured by mass cytometry. Values for immune cells correspond to frequency among all live cells. Points are colored by density; lines represent linear model fit with 95% confidence interval. See also **Figure S3**.

Somewhat expectedly, the frequency of total B cells and plasmablasts among CD45+ live cells increased with seroconversion (**Fig. 2a-c, Fig. S2a-c**). Other B cell subsets displaying significant positive association with seroconversion include key memory subsets such as switched Memory B cells (Switched Bmem), IgM+ memory B cells (IgM+ Bmem), c-delta switched memory B cells (c-delta switched B-mem), and pre-switched activated memory B cells (pre-switched activated Bmem) (**Fig. 2a-c, Fig. S2a-c**). Other B cell subsets enriched in seroconverted COVID19 patients include Mature naïve B cells and Age-dependent B cells (ABCs) (**Fig. 2a-b, Fig. S2a-c**). An increase in Mature naïve B cells is consistent with the development of humoral immunity. ABCs are associated with typical aging and development of autoimmunity, but their potential role during viral infections is less understood (13). Altogether, these findings illustrate the heterogenous immune state among hospitalized COVID19 patients, with seroconversion status being clearly associated with specific changes in circulating immune cell subsets, which could be largely understood as part of the progression of the antiviral immune response from innate cellular immunity to adaptive humoral immunity. As discussed later, these changes in immune cell frequencies occur in the context of clear lymphopenia, neutropenia, and thrombocytopenia in seronegative patients, with recovery of all these blood cell types in seropositive patients (see later, **Fig. 5-6**).

**Figure 4.**
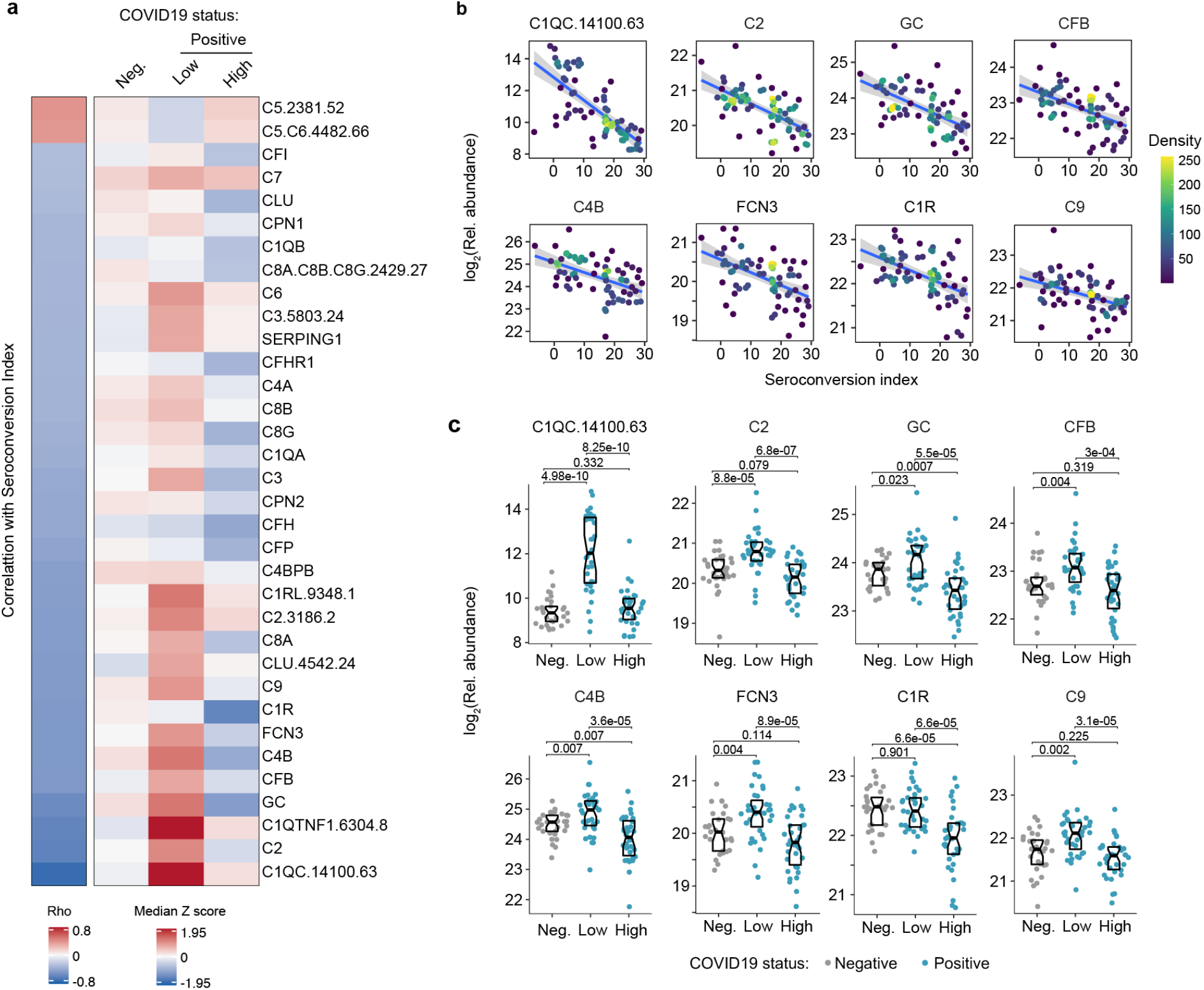
Seroconversion correlates with decreased markers of systemic complement activation. **a**. Heatmap displaying changes in circulating levels of components of the various complement pathways that are significantly correlated, either positively or negatively, with seroconversion status. The left column represents Spearman *rho* values, while the right columns display median Z-scores for each complement factor for controls (negative, Neg.) versus COVID19 patients (positive) divided into seroconversion low (Low) and high (High) status. Factors are ranked from most positively correlated (top, high *rho* values) to most anti-correlated (bottom, low *rho* values) with seroconversion status. **b**. Scatter plots for indicated complement factors significantly correlated with seroconversion indices among COVID19 patients. Points are colored by density; lines represent linear model fit with 95% confidence interval. **c**. Sina plots showing values for complement factors correlated with seroconversion comparing controls (Negative, Neg.) to COVID19 patients divided into seroconversion low (Low) and high (high) status. Data are presented as modified Sina plots with boxes indicating median and interquartile range. Numbers above brackets are Q-values for Mann-Whitney tests. See also **Figure S4**.

**Figure 5.**
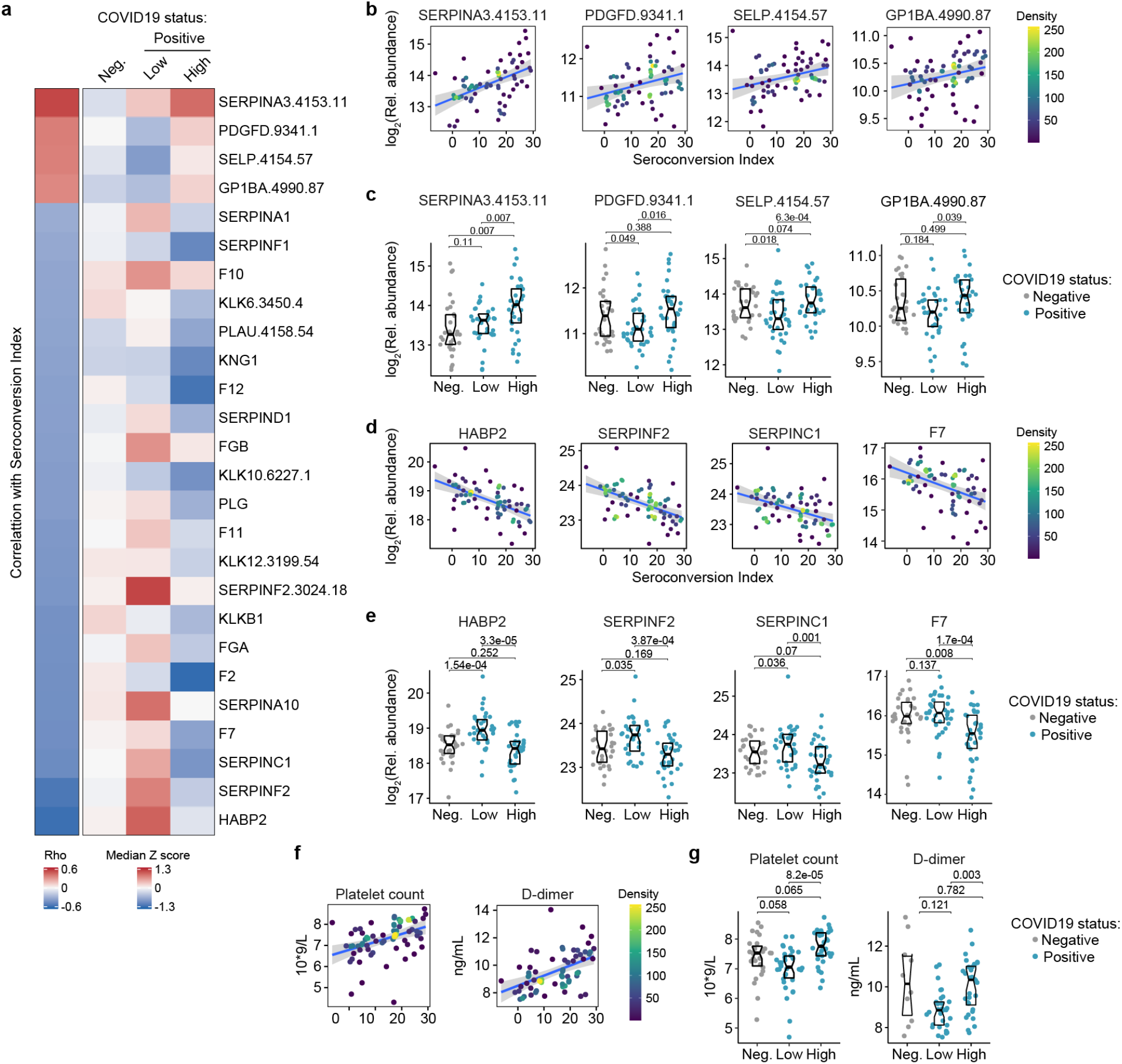
Seroconversion associates with remodeling of the hemostasis control network. **a**. Heatmap displaying changes in circulating levels of known modulators of hemostasis that are significantly correlated, either positively or negatively, with seroconversion status. The left column represents Spearman *rho* values, while the right columns display row-wise Z-scores for each factor for controls (negative, Neg.) versus COVID19 patients divided into seroconversion low (Low) and high (High) status. Factors are ranked from most positively correlated (top, high *rho* values) to most anti-correlated (bottom, low *rho* values) with seroconversion status. **b-c**. Scatter plots (b) and Sina plots (c) for factors positively correlated with seroconversion indices. **d-e**. Scatter plots (d) and Sina plots (e) for factors negatively correlated with seroconversion indices. **f-g**. Scatter plot (f) and Sina plots (g) displaying the correlations between seroconversion index and platelet counts and D-dimer values obtained from clinical laboratory testing. Points in (b), (d), (e) are colored by density; lines represent linear model fit with 95% confidence interval. Data in (c), (e), (g) are presented as modified Sina plots with boxes indicating median and interquartile range. Numbers above brackets are Q-values for Mann-Whitney tests. See also **Figure S5**.

**Figure 6.**
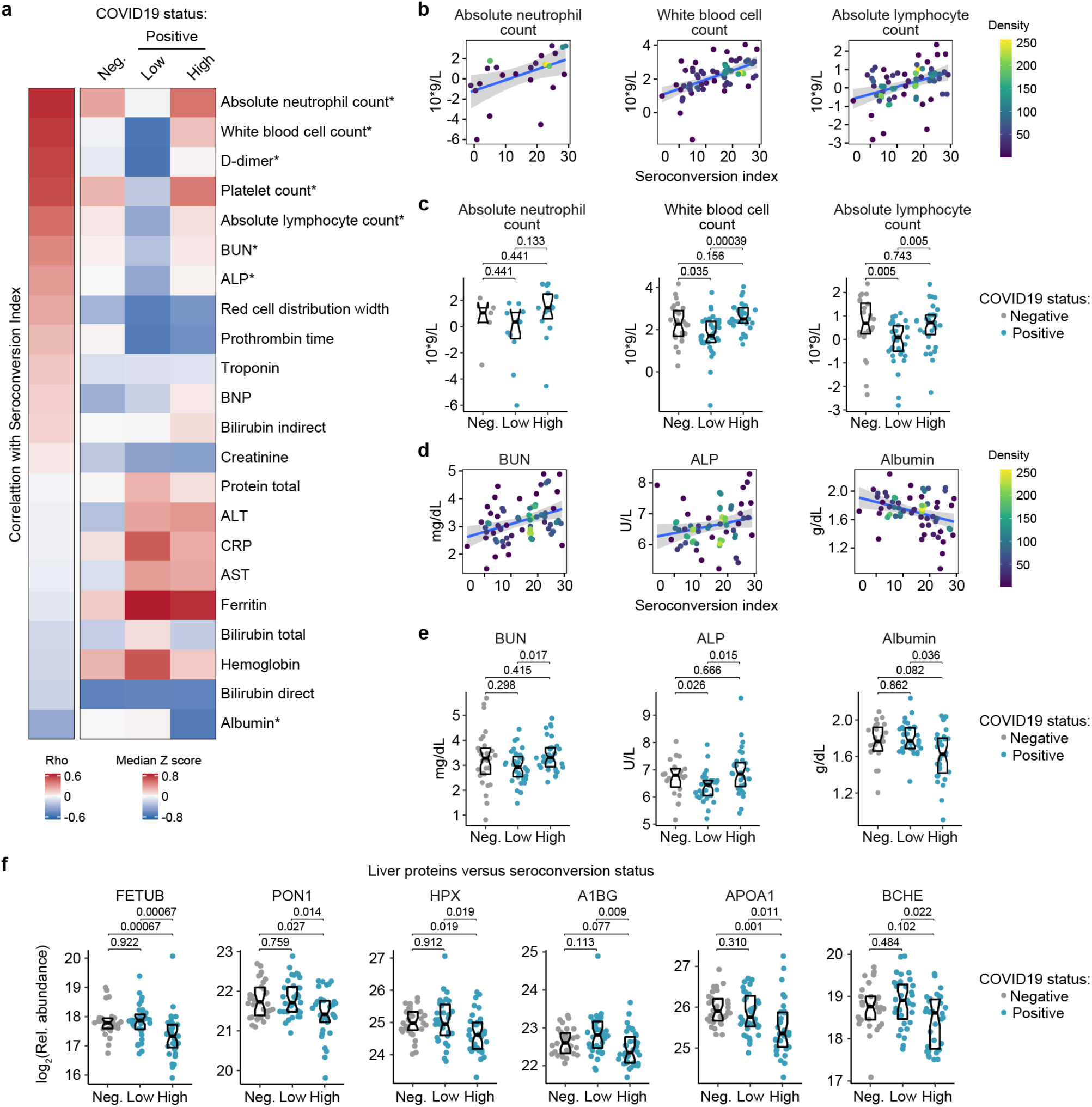
Seroconversion associates with recovery in blood cell numbers and hypoalbuminemia. **a**. Heatmap displaying correlations between clinical laboratory values and seroconversion status. The left column represents Spearman *rho* values, while the right columns display row-wise Z-scores for each variable for controls (negative, Neg.) versus COVID19 patients divided into seroconversion low (Low) and high (High) status. Measures are ranked from most positively correlated (top, high *rho* values) to most anti-correlated (bottom, low *rho* values) with seroconversion status. Asterisk indicates a significant correlation. **b-e**. Scatter plots (b and c) and Sina plots (c and e) for indicated clinical laboratory values significantly correlated with seroconversion indices among COVID19 patients. In b and c, points are colored by density; lines represent linear model fit with 95% confidence interval. In c and e, Sina plots show values for clinical laboratory tests correlated with seroconversion comparing controls (Negative, Neg.) to COVID19 patients divided into seroconversion low (Low) and high (High) status. Data are presented as modified Sina plots with boxes indicating median and interquartile range. Numbers above brackets are Q-values for Mann-Whitney tests. **f**. Sina plots showing values for liver proteins detected by MS that are significantly correlated with seroconversion comparing controls (Negative, Neg.) to COVID19 patients divided into seroconversion low (Low) and high (High) status. Data are presented as modified Sina plots with boxes indicating median and interquartile range. Numbers above brackets are Q-values for Mann-Whitney tests.

### Seroconversion associates with decreased IFN signaling

Next, we investigated associations between seroconversion and circulating levels of cytokines, chemokines, and other immune modulators in the bloodstream. Toward this end, we analyzed significant correlations in the MS proteomics, SOMAscan® proteomics, and MSD datasets (**Fig. S1b, Supp. Files S2-4**). Collectively, these three datasets contain data on dozens of factors involved in immune control (**Supp. File S6**). The most obvious result from this analysis was a clear negative correlation between seroconversion and circulating levels of key IFN ligands. Among 82 immune factors in the MSD dataset, top negative correlations are IFNA2, IFNL1, and IFNG (**Fig. S1b, Fig. 3a-b, Supp. Files S4** and **S6**). Among 5000+ epitopes measured by SOMAscan®, IFNA7, IFNL3 and IFNA1 rank among the top 10 negative correlations with seroconversion (**Fig. 3a-b, Supp. File S3**). All these IFN ligands were significantly higher in seronegative COVID19 patients relative to the control cohort, but levels fall back within normal ranges in seropositive COVID19 patients (**Fig. 3a,c**). These results could be interpreted as a transient wave of IFN production during early stages of SARS-CoV-2 infection, with return to normal levels upon development of humoral immunity. This notion is further supported by the behavior of key IFN-inducible proteins, such as CXCL10 (C-X-C Motif Chemokine Ligand 10, IFN-inducible protein 10, IP10) (**Fig. 3a, Fig. S3a-b**). Notably, this pattern was not evident for IFNB1 (**Fig. S3a-b**). Factors involved in monocyte differentiation and activation were also preferentially elevated in seronegative patients, such as CCL8 (C-C Motif Chemokine Ligand 8, Monocyte Chemoattractant Protein 2, MCP2), CSF3 (Colony Stimulating Factor 3, Granulocyte Colony Stimulating Factor, G-CSF), and CCL19 (C-C Motif Chemokine Ligand 19, Macrophage Inflammatory Protein 3 beta, MIP3beta) (**Fig. 3a-c**). Although circulating levels of total monocytes and monocyte subsets are not significantly correlated with seroconversion status (**Supp. File S5**), these results are consistent with a transient round of activation and mobilization of tissue-resident monocytes and macrophages by local IFN production, with subsequent decreases upon seroconversion. In support of this notion, we noticed that circulating levels of CD14, a surface marker for monocytes and macrophages, were strongly anticorrelated with seroconversion, being significantly elevated among seronegative patients and significantly depleted in seropositive patients (**Fig. S3a-b**). In fact, CD14 was the top negative correlation in the MS proteomics dataset (**Fig. S1b, Supp. File S2**).

In agreement with the signs of B cell maturation and differentiation associated with seroconversion (**Fig. 2a-b** and **S2a-c**), top correlations among immune factors include TNFSF13B (TNF Superfamily Member 13B, B-cell activating factor, BAFF), and its receptor, TNFRSF17 (TNF Receptor Superfamily Member 17, B cell Maturation Protein, BCMA). TNFSF13B is increased preferentially in seronegative patients relative to the control group (**Fig. 3c**). In contrast, its receptor TNFRSF17 decreases preferentially in seronegative patients, returning to levels similar to the control group upon seroconversion (**Fig. 3c**). The increased levels of TNFSF13B in seronegative patients are consistent with a strong wave of B cell stimulation and proliferation prior to B cell maturation and seroconversion. The decrease in circulating soluble TNFRSF17 could be interpreted as a consequence of transient lymphopenia prior to seroconversion (see later **Fig. 6**). Other interesting top correlations reveal that seroconversion associates with a restoration of circulating cytokines depleted preferentially in seronegative COVID19 patients, such as CCL14 and CCL24 (Eotaxin-2) (**Fig. 3a**). Again, these changes could be explained by decreases in lymphocyte counts preferentially in seronegative patients (see later, **Fig. 6**). Of note, seroconversion is not strongly correlated with changes in acute phase proteins (e.g. C-reactive protein, ferritin, **Supp. Files S2-4**). Whereas CRP levels measured by MS decrease somewhat in seronegative patients, ferritin levels remain high (**Fig. S3c**), suggesting that seroconversion does not fully reverse the broader inflammatory phenotype of COVID19.

In order to understand how these changes in cytokines could be integrated with changes observed in circulating immune cell types in the MC dataset, we interrogated whether levels of IFNA2, the top anticorrelated cytokine with seroconversion indices, showed significant correlations with immune cell subsets among all live peripheral blood mononuclear cells (PBMCs) (**Supp. File S7**). Indeed, IFNA2 levels correlated negatively with key B cell subsets increased upon seroconversion, and positively with pDCs, T cell subsets decreased upon seroconversion, and CD56^bright^ NK cells (**Fig. 3d, Fig. S3d, Supp. File S7**).

Altogether, these observations could be interpreted as an orchestrated movement in the immune system away from an innate immune response marked by IFN production and IFN-inducible changes in immune cell type frequency and function, toward a state of adaptive humoral immunity and antibody production.

### Seroconversion associates with decreased markers of systemic complement activation

Analysis of the top negative correlations with the MS and SOMAscan^®^ proteomics datasets revealed that seroconversion correlates strongly with decreased plasma levels of subunits of the various complement pathways (**Supp. Files S2-3**). In fact, 10 of the top 20 negative correlations in the MS dataset are complement subunits or complement regulators, and the top negative correlation in the SOMAscan® dataset is the complement subunit C1QC (**Fig. 4a-b, Fig. S1b-c, Supp. Files 2-3**). This led us to complete a more thorough investigation of the interplay between seroconversion and the complement pathways (**Supp. File S8**).

There are three recognized complement pathways, known as the classical, lectin, and alternative pathways, with significant crosstalk among them and convergence on the so called terminal pathway that leads to formation of the membrane attack complex (MAC) (14). Proteins from all three pathways were significantly anti-correlated with seroconversion including C1QA, C1QB, C1QC, C1R, and C1S, all involved in initiation of the classical pathway; C2, C4A, and C4B, which share functions in activation of the classical and lectin pathways; C3, which acts both in the lectin and alternative pathways; as well as C6, C7, C8A, C8B, C8G, and C9, which act in the downstream terminal pathway (**Fig. 4a**). Additionally, seroconversion correlates negatively with positive regulators of the complement cascade, such as GC (GC Vitamin D Binding Protein), which enhances the chemotactic activity of C5 alpha for neutrophils in inflammation and macrophage activation (15); FCN3 (Ficolin 3), a protein involved in activation of the lectin complement pathway (16); CFB (Complement Factor B, C3/C5 Convertase) and CFP (Complement Factor P, Properdin). Most of these factors are significantly elevated in seronegative COVID19 patients relative to controls, but return to baseline or below baseline levels in seronegative patients (**Fig. 4c** and **S2a-b**). Negative modulators of complement function showed similar behaviors, such as CFH (Complement Factor H), C4BPB (C4b binding protein) and SERPING1 (C1 inhibitor), suggesting the induction of negative feedback mechanisms during complement activation in seronegative patients (**Fig. 4a**). Only SOMAscan® signals for C5 and the C5.C6 complex showed the opposite behavior, with lower signals in seronegative COVID19 patients, which could be interpreted as increased consumption of the C5 precursor polypeptide by the C5 convertase (**Fig. 4a, Fig. S4a**). Altogether, although our proteomics platforms do not enable a complete characterization of the complement cascade in terms of measuring cleaved fragments, protein complexes, and post-translational modifications, our results can nonetheless be understood as a profound, yet transient wave of systemic activation of the complement cascades early during the course of SARS-CoV-2 infections, followed by return to normal levels upon seroconversion.

### Seroconversion associates with remodeling of the hemostasis network toward platelet recovery and activation

Top positive and negative correlations between seroconversion indices and the proteomics datasets included many prominent regulators of hemostasis (**Supp. Files S2-3)**. Given the importance of thromboembolism and microangiopathies in COVID19, we decided to investigate the interplay between seroconversion and circulating levels of factors involved in coagulation and thrombosis in more detail (**Supp. File S9**). Among these factors, positive correlations with seroconversion indices were dominated by markers of platelet degranulation (**Fig. 5a**), including key proteins stored in platelet alpha granules, such as SERPINA3 (Alpha-1-antichymotrypsin, ACT), PDGFD (Platelet Derived Growth Factor D), SELP (Selectin P), and GP1BA (glycoprotein Ib alpha) (**Fig. 5a-c, Supp. File S9**). Except for SERPINA3, all factors are depleted in seronegative patients relative to the control cohort, and all four factors are significantly elevated in seroconverted patients relative to seronegative patients (**Fig. 5a,c**). Conversely, top negative correlations with seroconversion in both proteomics datasets include key modulators of the intrinsic and extrinsic coagulation pathways. Many positive regulators of coagulation are elevated in seronegative patients and depleted in seropositive patients, including HABP2 (hyaluronan activated binding protein 2, factor VII activating protein, FSAP), SERPINF2 (alpha-2-plasmin inhibitor), F7 (Coagulation Factor VII), F2 (Coagulation Factor II, thrombin), F11 (Coagulation Factor XI), F12 (Coagulation Factor XII), and F10 (Coagulation Factor X), as well as the structural components FGA (Fibrinogen Alpha Chain) and FGB (Fibrinogen Beta Chain) (**Fig. 5a, d-e, Fig. S5a-b**). However, many endogenous anticoagulants and drivers of fibrinolysis also show a similar pattern, such as SERPINC1 (antithrombin, AT-III), SERPINA10 (antitrypsin), PLG (Plasminogen), and PLAU (Plasminogen Activator, Urokinase). SERPINC1 is a potent inhibitor of thrombin, as well as coagulation factors IXa, Xa and XIa (17). SERPIN10A is another inhibitor of coagulation factors Xa and XIa (17). Plasminogen is the precursor of plasmin, the key enzyme in fibrinolysis, and PLAU mediates proteolytic generation of plasmin. Lastly, several kallikreins, including KLKB1, KLK12, KLK10 and KLK12, as well as KNG1 (kininogen) are all depleted in seroconverted patients (**Fig. S5a-b**). The kinin-kallikrein system plays key roles in coagulation, inflammation, and blood pressure control (18). Once activated, kallikreins function as serine proteases that can cleave plasminogen into plasmin, thus promoting fibrinolysis, but also high-molecular weight kininogen (HMWK) into the vasoactive peptide bradykinin, thus promoting vasodilation (18).

Overall, the interpretation of varying plasma levels for these various modulators of hemostasis is not straightforward, as many of these factors are subject to proteolytic cleavage, consumption, and/or aggregation, and many of them are produced by the liver and cleared by the kidney, two organs affected in COVID19. Therefore, in order to place these complex proteomic signatures in the context of COVID19 pathology, we investigated correlations between seroconversion indices and clinical laboratory values for platelets and D-dimer measured in the course of hospitalization. As part of standard of care in hospitalized COVID19 patients, platelets are routinely counted to assess thrombocytopenia, whereas D-dimer, a proteolytic product of blood clots during fibrinolysis, is routinely measured to assess thrombotic risk. We therefore obtained platelet counts and D-dimer values from the clinical laboratory tests closest in time to the research blood draw employed for the -omics measurements. Both platelet counts and D-dimer correlated positively with the seroconversion scores (**Fig. 5f**), being depleted in seronegative patients relative to the control group and significantly higher in seropositive patients relative to seronegative patients (**Fig. 5g**).

Altogether, these results are consistent with transient thrombocytopenia early in the course of SARS-CoV-2 infections, followed by recovery in platelet counts, increased platelet degranulation, and higher levels of fibrinolysis products in seroconverted patients. Additionally, as discussed next, depletion of coagulation factors produced by the liver in seropositive patients could be tied to liver dysfunction and/or vascular leakage.

### Seroconversion is accompanied by emergency hematopoiesis and hypoalbuminemia

Analysis of the clinical laboratory values obtained closest in time to the research blood draws revealed other significant correlations between key clinical parameters and seroconversion indices. In addition to the aforementioned positive correlations with platelet counts and D-dimer levels, seroconversion correlated positively and significantly with absolute neutrophil count, white blood cell count, and absolute lymphocyte count (**Fig. 6a-c, Fig. S6a, Supp. File S10**). Consistently, these parameters are lower in the seronegative COVID19 patients, but return to baseline levels in seroconverted patients. These changes are indicative of an early but transient round of neutropenia, thrombocytopenia and lymphopenia in COVID19, followed by ‘emergency hematopoiesis’, a compensatory phenomenon involving broad stimulation of hematopoietic and stem cell progenitors (HSPCs) by factors such as G-CSF (CSF3) (19), which we found to be elevated in seronegative patients (**Fig. 3a**) The oscillations in neutrophils and lymphocyte counts could explain some of the changes in soluble immune factors depleted in seronegative patients only (e.g. TFNFRSF17 expressed by B cells) (**Fig. 3a-c**).

Other significant correlations revealed that seroconversion associates with markers of liver dysfunction and/or vascular leakage. Seroconverted patients showed elevated levels of blood urea nitrogen (BUN), elevated alkaline phosphatase (ALP), and decreased levels of albumin (ALB) (**Fig. 6a, d-e**). Elevated BUN is indicative of liver and/or kidney malfunction. Elevated ALP is indicative of liver damage.

Although the markers of liver injury AST (aspartate transaminase) and ALT (alanine transaminase) are elevated in the COVID19 cohort relative to the control group, they are not significantly associated with seroconversion status (**Fig. 6a, Fig. S6a-b**). Creatinine was significantly lower in COVID19 patients, also a sign of liver pathology, but no different by seroconversion status (**Fig. S6a,b**). Low levels of circulating albumin, of hypoalbuminemia, is a common feature of COVID19 pathology that has been associated with worse prognosis independently of age and comorbidities (20). It has been proposed that marked hypoalbuminemia without differences in AST and ALT could be due to inflammation-driven escape of serum albumin into interstitial space downstream of increased vascular permeability (20). In order to investigate this further, we probed the MS proteomics dataset to see if other abundant liver-derived proteins showed similar behavior (**Supp. File S2**). Indeed, seroconversion correlates with significantly decreased levels of major liver-derived proteins such as FETUB (fetuin B), PON1 (Paraoxonase 1), HPX (hemopexin), A1BG (Alpha-1b-glycoprotein), APOA1 (apolipoprotein A1), and BCHE (Butyrylcholinesterase) (**Fig. 6f, Supp. File S2**). This phenomenon could explain why many liver-derived factors involved in hemostasis control are also depleted in seropositive patients (e.g. fibrinogens, F2, F7, kallikreins) (**Fig. 5a, Fig. S5a-b**).

Altogether, these results reveal that seroconversion is associated with recovery in diverse white blood cell types, indicative of emergency hematopoiesis, along with biomarkers indicative of more severe liver dysfunction and/or increased vascular damage.

## DISCUSSION

The temporal sequence of seroconversion relative to the onset of COVID19 symptoms has been already established (21, 22). Within two weeks of symptom onset, virus-specific antibodies start accumulating in the bloodstream (22). Circulating IgMs, IgAs and IgGs against SARS-CoV-2 increase rapidly thereafter, followed by decay of IgMs and IgAs over time, while IgGs remain high for several weeks and months (22). Building upon this knowledge, our integrated analysis of biosignatures of seroconversion using IgG measurements against SARS-CoV-2 polypeptides in hospitalized COVID19 patients supports a model for staging COVID19 pathology into a distinct sequence of early and late events, referred hereto as Stage 1 and Stage 2 (**Fig. 7**).

**Figure 7.**
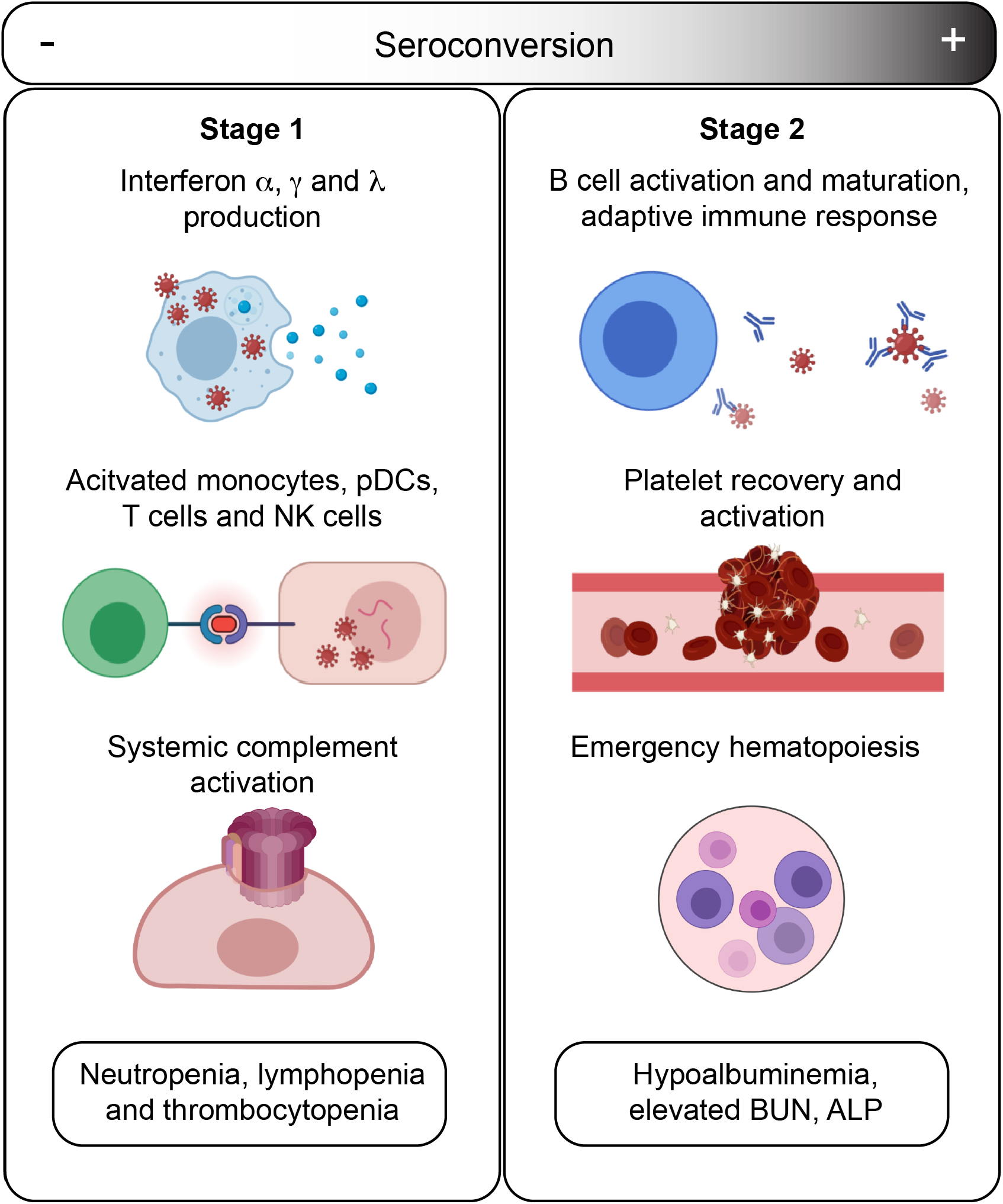
Model for staging of COVID19 pathological process based on seroconversion status. Stage 1 applies to COVID19 patients with low degree of seroconversion and involves high levels of circulating IFNs, signs of strong systemic complement activation, hyperactive T cells, activated monocytes and cytokine-producing NK cells, as well as diverse cytopenias. Stage 2 applies to COVID19 patients with high degree of seroconversion, and is characterized by recovered blood cellnumbers, increased levels of markers of platelet degranulation, increased liver dysfunction and/or hypoalbuminemia.

In Stage 1, associated with the initial antiviral response, hospitalized patients carry low levels of SARS-CoV-2-specific IgGs, high levels of circulating IFN ligands and complement subunits, signatures of activated T cells, pDCs and monocytes, high levels of cytokine-producing NK cells, as well as neutropenia, thrombocytopenia, and lymphopenia. In Stage 2, associated with development of humoral immunity, patients carry high levels of SARS-CoV-2-specific antibodies, near baseline levels of IFN ligands, complement subunits, and activated T cells, no significant cytopenias, but clear signs of B cell differentiation, plasmablast accumulation, platelet degranulation markers, and increased markers of liver damage (ALP, BUN) and hypoalbuminemia.

Based on our findings, we propose the following sequence of events in mild-to-moderate symptomatic, hospitalized COVID19 patients. In Stage 1, COVID19 pathology is dominated by IFN- and complement-driven processes. Although SARS coronaviruses have evolved diverse strategies to evade the antiviral effects of IFNs (23), IFN signaling nonetheless remains a potent defense mechanism against SARS-CoV-2, as illustrated by the fact that both genetic variants compromising IFN signaling and autoantibodies against IFN ligands have been associated with severe COVID19 (24, 25). However, sustained high levels of Type I and Type III IFN ligands could potentially contribute to SARS pathology through disruption of lung barrier function and other mechanisms (6, 7). In mouse models of both SARS-CoV-1 and SARS-CoV-2 infections, Type I IFN signaling was shown to be required for development of lung pathology (26, 27). In Stage 1, high IFN signaling is likely to drive activation of monocytes and T cells as well as B cell differentiation. In our studies, levels of IFNA2 are clearly correlated with activation of both CD4+ and CD8+ T cell subsets, with polarization of CD4+ T cells toward Th1 and Th17 states. Stage 1 is also correlated with signs of monocyte activation, demarked by high levels of MCP2/CCL8, CSF3, and circulating CD14, as well as high production of TNFSF13B. Stage 1 is also characterized by high levels of circulating complement factors. Recent studies have shown direct activation of the alternative complement pathway by the SARS-CoV-2 spike protein (28), and systemic complement activation has been associated with severe respiratory failure in COVID19 (8, 29). Sustained high levels of complement activation could contribute to pathology through both the cytolytic effects of the MAC and the proinflammatory effects of the C5a-C5aR1 axis (29). Indeed, in a mouse model of SARS-CoV-1, depletion of the C3 complement subunit attenuated SARS pathology (30), and anti-C5aR1 neutralizing antibodies inhibited the C5a-mediated recruitment of human myeloid cells to lung tissue and reduced accompanying lung injury in a humanized mouse model (29).

Complement could damage the endothelial tissue in the lung and other organs, compromising vascular integrity. Stage 1 is also characterized by neutropenia, lymphopenia, and thrombocytopenia.

In this model, the transition from Stage 1 to Stage 2 is mediated by development of humoral immunity, as well as emergency hematopoiesis. As B cells differentiate, mature, and are selected by clonal evolution, plasmablasts are enriched in the bloodstream, producing specific IgGs of ever greater neutralizing capacity recognizing the RBD region of the spike protein. Seroconversion not only prevents viral reentry into cells, but could also prevent direct activation of the complement cascade by the spike protein (28). As humoral immunity develops, levels of IFN signaling and complement activity plummet. Increased levels of broad stimulators of hematopoiesis in Stage 1 (e.g. CSF3) would lead to emergency hematopoiesis. In turn, increased numbers of circulating platelets during Stage 2 would encounter the endothelial damage caused during Stage 1 by the cytolytic effects of the virus, disruption of lung barrier function by IFNs, complement attack, and even perhaps T cell- and monocyte-mediated cellular toxicity. In turn, the vascular damage ensued during Stage 1 would contribute to leakage of serum proteins in Stage 2 (i.e. hypoalbuminemia). Although our analyses revealed significant remodeling of the hemostasis network in Stage 1 and Stage 2, we can not conclusively interpret these results in terms of relative risk of thromboembolism and microangiopathy. Although D-dimer levels are higher in seropositive patients, our study is not powered to conclude if thrombotic disease is higher upon seroconversion.

According to this model, therapeutic interventions being tested in COVID19 could benefit from patient stratification based on a quantitative assessment of seroconversion. For example, JAK inhibitors and complement inhibitors may be more effective in cohorts enriched for seronegative patients. Clinical biomarkers for Stage 1 could be high IFN levels, high complement levels, and/or severe cytopenia. In contrast, patients in Stage 2 may not benefit from JAK inhibitors and complement inhibitors. Instead, they could benefit from better management of liver dysfunction and/or vascular damage. Notably, albumin supplementation was found to improve oxygenation in ARDS (31). This model also indicates that analysis of clinical trial data for various therapeutic interventions should take into consideration seroconversion status, as results could vary between Stage 1 and Stage 2 patients.

Given the cross-sectional nature of our study, our model should be challenged with longitudinal analysis of the biosignatures reported here. Out study does not address differences between patients with mild, moderate, and severe disease, and our data does not include analysis of patients admitted to ICU with life-threatening COVID19. Nevertheless, our model indicates that time span between exposure to virus and seroconversion could be a key determinant of disease severity. The longer an affected individual remains in Stage 1, the more likely that the potential harmful effects of IFN hyperactivity and complement toxicity will be manifested, with increased likelihood of endothelial and organ damage, thus setting the stage for more severe thrombotic events and vascular damage in Stage 2. Notably, markers of disease severity could be found in either stage. For example, high IFNA2 (Stage 1 marker) and strong hypoalbuminemia (Stage 2 marker), have been independently associated with risk of severe COVID19 (20, 34). A delay in seroconversion could potentially explain in part the high risk of severe COVID19 pathology in the elder, as B cell function and development of humoral immunity are decreased with age (33).

In sum, our results support the existence of distinct pathophysiological states among hospitalized COVID19 patients, with seroconversion status being potentially useful as a surrogate marker of underlying processes. We hope these results will prompt additional investigations into the sequence of pathological events in COVID19 and how to ameliorate them for therapeutic purposes.

## METHODS

### Study design, participant recruitment, and clinical data capture

Research participants were recruited and consented for participation in the COVID Biobank of the University of Colorado Anschutz Medical Campus [Colorado Multiple Institutional Review Board (COMIRB) Protocol # 20-0685]. Data was generated from deidentified biospecimens and linked to demographics and clinical metadata procured through the Health Data Compass of the University of Colorado under COMIRB Protocol # 20-1700. Participants were hospitalized either at Children’s Hospital Colorado or the University of Colorado Hospital. COVID status was defined by a positive PCR reaction and/or antibody test. Cohort characteristics can be found in **Supp. File S1**.

### Blood processing

Blood samples were collected into EDTA tubes and sodium heparin tubes. After centrifugation, EDTA plasma was used for MS proteomics, SOMAscan^®^ proteomics, as well as multiplex immunoassays using MSD technology for both cytokine profiles and seroconversion assays. From sodium heparin tubes, PBMCs were obtained by the Ficoll gradient method before cryopreservation and assembly of batches for MC analysis (see below).

### Plasma proteomics by mass spectrometry

Plasma samples were digested in S-Trap filters (Protifi, Huntington, NY) according to the manufacturer’s procedure. Briefly, a dried protein pellet prepared from organic extraction of patient plasma was solubilized in 400 µl of 5% (w/v) SDS. Samples were reduced with 10 mM DTT at 55°C for 30 min, cooled to room temperature, and then alkylated with 25 mM iodoacetamide in the dark for 30 min. Next, a final concentration of 1.2% phosphoric acid and then six volumes of binding buffer [90% methanol; 100 mM triethylammonium bicarbonate (TEAB); pH 7.1] were added to each sample. After gentle mixing, the protein solution was loaded into an S-Trap filter, spun at 2000 rpm for 1 min, and the flow-through collected and reloaded onto the filter. This step was repeated three times, and then the filter was washed with 200 μL of binding buffer 3 times. Finally, 1 μg of sequencing-grade trypsin (Promega) and 150 μL of digestion buffer (50 mM TEAB) were added onto the filter and digestion carried out at 47 °C for 1 h. To elute peptides, three stepwise buffers were applied, 200 μL of each with one more repeat, including 50 mM TEAB, 0.2% formic acid in H_2_O, and 50% acetonitrile and 0.2% formic acid in H_2_O. The peptide solutions were pooled, lyophilized and resuspended in 1 mL of 0.1 % FA. 20 µl of each sample was loaded onto individual Evotips for desalting and then washed with 20 μL 0.1% FA followed by the addition of 100 μL storage solvent (0.1% FA) to keep the Evotips wet until analysis. The Evosep One system (Evosep, Odense, Denmark) was used to separate peptides on a Pepsep column, (150 µm internal diameter, 15 cm) packed with ReproSil C18 1.9 µm, 120A resin. The system was coupled to a timsTOF Pro mass spectrometer (Bruker Daltonics, Bremen, Germany) via a nano-electrospray ion source (Captive Spray, Bruker Daltonics). The mass spectrometer was operated in PASEF mode. The ramp time was set to 100 ms and 10 PASEF MS/MS scans per topN acquisition cycle were acquired. MS and MS/MS spectra were recorded from *m/z* 100 to 1700. The ion mobility was scanned from 0.7 to 1.50 Vs/cm^2^. Precursors for data-dependent acquisition were isolated within ± 1 Th and fragmented with an ion mobility-dependent collision energy, which was linearly increased from 20 to 59 eV in positive mode. Low-abundance precursor ions with an intensity above a threshold of 500 counts but below a target value of 20000 counts were repeatedly scheduled and otherwise dynamically excluded for 0.4 min. Raw data file conversion to peak lists in the MGF format, downstream identification, validation, filtering and quantification were managed using FragPipe version 13.0. MSFragger version 3.0 was used for database searches against a Human isoform-containing UniProt fasta file (version 08/11/2020) with decoys and common contaminants added. The identification settings were as follows: Trypsin, Specific, with a maximum of 2 missed cleavages, up to 2 isotope errors in precursor selection allowed for, 10.0 ppm as MS1 and 20.0 ppm as MS2 tolerances; fixed modifications: Carbamidomethylation of C (+57.021464 Da), variable modifications: Oxidation of M (+15.994915 Da), Acetylation of protein N-term (+42.010565 Da), Pyrolidone from peptide N-term Q or C (−17.026549 Da). The Philosopher toolkit version 3.2.9 (build 1593192429) was used for filtering of results at the peptide and protein level at 0.01 FDR. Label-free quantification was performed by AUC integration with matching between all runs using IonQuant.

### Plasma proteomics by SOMAscan^®^ assays

125 μL EDTA plasma was analyzed by SOMAscan^®^ assays using previously established protocols (35). Briefly, each of the 5000+ SOMAmer reagents binds a target peptide and is quantified on a custom Agilent hybridization chip. Normalization and calibration were performed according to SOMAscan^®^ Data Standardization and File Specification Technical Note (SSM-020) (35). The output of the SOMAscan^®^ assay is reported in relative fluorescent units (RFU).

### Cytokine profiling and seroconversion by multiplex immunoassay

Multiplex immunoassays MSD assays were performed on EDTA plasma aliquots following manufacturer’s instructions (Meso Scale Discovery, MSD). A list of immune factors measured by MSD can be found in **Supp. File S4**. Values were extrapolated against a standard curve using provided calibrators. Seroconversion assays against SARS-CoV-2 proteins and the control protein from the Flu A Hong Kong H3 virus were performed in a multiplex immunoassay using the IgG detection readout according to manufacturer’s instructions (MSD). Relative values were extrapolated against a standardized curve consisting of pooled COVID19 positive reference plasma (36).

### Mass cytometry analysis of immune cell types

Cryopreserved PBMCs were thawed, washed twice with Cell Staining Buffer (CSB) (Fluidigm), and counted with an automated cell counter (Countess II - Thermo Fisher Scientific). Extracellular staining on live cells was done in CSB for 30 min at room temperature, in 3-5^10^6^ cells per sample. Cells were washed with 1X PBS (Fluidigm) and stained with 1 mL of 0.25 mM cisplatin (Fluidigm) for 1 min at room temperature for exclusion of dead cells. Samples were then washed with CSB and incubated with 1.6% PFA (Electron Microscopy Sciences) during 10 min at room temperature. Samples were washed with CBS and barcoded using a Cell-IDTM 20-Plex Pd Barcoding Kit (Fluidigm) of lanthanide-tagged cell reactive metal chelators that will covalently label samples with a unique combination of palladium isotopes, then combined. Surface staining with antibodies that work on fixed epitopes was performed in CSB for 30 min at room temperature (see **Supp. File S11** for antibody information). Cells were washed twice with CSB and fixed in Fix/Perm buffer (eBioscience) for 30 min, washed in permeabilization buffer (eBioscience) twice, then intracellular factors were stained in permeabilization buffer for 45 min at 4°C. Cells were washed twice with Fix/Perm Buffer and were labeled overnight at 4°C with Cell-ID Intercalator-Ir (Fluidigm) for DNA staining. Cells were then analyzed on a Helios instrument (Fluidigm). To make all samples comparable, pre-processing of mass cytometry data included normalization within and between batches via polystyrene beads embedded with lanthanides as previously described (37). Files were debarcoded using the Matlab DebarcoderTool (38). Then normalization again between batches relative to a reference batch based on technical replicates (39). Gating was performed using CellEngine (Primitybio). Gating strategy is summarized in **Fig. S1c-g** and **Supp. File S12**.

### Biostatistics and bioinformatics analyses

Preprocessing, statistical analysis, and plot generation for all datasets was carried out using R (R 4.0.1 / Rstudio 1.3.959 / Bioconductor v 3.11) (40-42), as detailed below. *MSD seroconversion data*. Plasma concentration values (pg/mL) for IgGs recognizing SARS-Co-V-2 and Flu A Hong Kong H3 epitopes were adjusted for Sex and Age using the *removeBatchEffect* function from the limma package (v 3.44.3) (43). Distributions of Sex/Age-adjusted concentration values for each epitope in COVID19 positive and COVID19 negative samples were compared using the Wilcoxon-Mann-Whitney two-sample rank-sum test, with Benjamini-Hochberg correction of p-values and an estimated false discovery rate threshold of 0.1. To capture seroconversion as a single value we calculated a ‘seroconversion index’ for each sample as follows. Firstly, Z-scores were calculated from the adjusted concentration values for each epitope in each sample, based on the mean and standard deviation of COVID19-negative samples. Secondly, the per-sample seroconversion index was calculated as the sum of Z-scores for the four SARS-CoV-2 seroconversion assays. For comparison of multiple measurements from COVID19 positive samples with high seroconversion indices to those with low seroconversion indices, or COVID19 negative samples, COVID19 positive samples were divided into two equal-sized groups based on their seroconversion index, referred to as ‘seronegative’ versus ‘seropositive’ groups.

#### MS-proteomic data

Raw Razor intensity data were filtered for high abundance proteins by removing those with >70% zero values in both COVID19 negative and COVID19 positive groups. For the remaining 407 abundant proteins, 0 values (8,363 missing values of 44,363 total measurements) were replaced with a random value sampled from between 0 and 0.5x the minimum non-zero intensity value for that protein. Data was then normalized using a scaling factor derived from the global median intensity value across all proteins / sample median intensity across all proteins (44) and adjusted for Sex and Age using the *removeBatchEffect* function from the limma package (v 3.44.3) (43).

#### SOMAscan^®^ data

Normalized data (RFU) was imported and converted from a SOMAscan^®^ .adat file using a custom R package (SomaDataIO) and adjusted for Sex and Age using the *removeBatchEffect* function from the limma package (v 3.44.3) (43).

#### MSD cytokine profiling data

Plasma concentration values (pg/mL) for each of the cytokines and related immune factors measured across multiple MSD assay plates was imported to R, combined, and analytes with >10% of values outside of detection or fit curve range flagged. For each analyte, missing values were replaced with either the minimum (if below fit curve range) or maximum (if above fit curve range) calculated concentration and means of duplicate wells used in all further analysis. Data was adjusted for Sex and Age using the *removeBatchEffect* function from the limma package (v 3.44.3) (43).

#### Analysis of mass cytometry data

Cell population frequencies, exported from CellEngine as percentages of various parental lineages, were adjusted for Sex and Age using the *removeBatchEffect* function from the limma package (v 3.44.3) (43).

#### Correlation analysis

To identify features in each dataset that correlate with Seroconversion Index in COVID19 positive samples, Spearman *rho* values and p-values were calculated against the Sex/Age-adjusted values for each dataset using the *rcorr* function from the Hmisc package (v 4.4-0) (45), with Benjamini-Hochberg correction of p-values and an estimated false discovery fate threshold of 0.1. For visualization, XY scatter plots with points colored by local density were generated using a custom density function and the ggplot2 (v3.3.1) package (46).

#### Comparison of seroconversion groups

Distributions of Sex/Age-adjusted concentration values for features in COVID19 negative samples and COVID19 positive samples with low vs. high Seroconversion Indices were compared with pair-wise Wilcoxon-Mann-Whitney two-sample rank-sum tests, using the *wilcox_test* function from the rstatix package (v0.6.0) (47), with Benjamini-Hochberg correction of p-values and an estimated false discovery rate threshold of 0.1. To visualize differences between COVID negative samples and COVID positive samples with low vs. high Seroconversion Indices, Z-scores were calculated for each feature based on the mean and standard deviation of COVID negative samples, and visualized as heatmaps and/or modified sina plots using the ComplexHeatmap (v2.4.2) (48), ggplot2 (v3.3.1), and ggforce (v0.3.1) packages (49).

## Data Availability

All data generated for this manuscript is made available through the online researcher gateway of the COVIDome Project, known as the COVIDome Explorer, which can be accessed at covidome.org. The mass spectrometry proteomics data have been deposited to the ProteomeXchange Consortium via the PRIDE partner repository with the dataset identifier PXD022817. The mass cytometry data has been deposited in Flow Repository under the link: https://flowrepository.org/id/RvFrPyf61bmFlccxkki30pKSU83yWnVEky943X4winAnC7xDP1b7Lu0G36zhv3Qh. The SOMAscan Proteomics, MSD Cytokine Profiles, and Sample Metadata files have been deposited in Mendeley under entry doi:10.17632/2mc6rrc5j3.1.

https://medschool.cuanschutz.edu/covidome/

## DATA AVAILABILITY STATEMENT

All data generated for this manuscript is made available through the online researcher gateway of the COVIDome Project, known as the COVIDome Explorer, which can be accessed at covidome.org. The mass spectrometry proteomics data have been deposited to the ProteomeXchange Consortium via the PRIDE partner repository (50) with the dataset identifier PXD022817. The mass cytometry data has been deposited in Flow Repository under the link: https://flowrepository.org/id/RvFrPyf61bmFlccxkki30pKSU83yWnVEky943x4winAnC7xDP1b7Lu0G36zhv3Qh. The SOMAscan® Proteomics, MSD Cytokine Profiles, and Sample Metadata files have been deposited in Mendeley under entry doi:10.17632/2mc6rrc5j3.1.

## AKNOWLEDGMENTS

We are grateful to Dr. Thomas Flaig for his leadership in setting up the COVID19 Biobank at the University of Colorado and also to the COVID19 Biobank Steering Committee for overall support of this project. We thank members of the Biorepository Shared Resource, especially Dr. Adrie Van Bokhoven, Zachary Grasmick, and Hannah Schumman, and members of the Human Immune Monitoring Shared Resource, especially Dr. Jill Slansky, Jodi Livesay, Troy Schedin, and Jennifer McWilliams, as well as Aaron Issaian for assistance with MS proteomics data analysis. We also thank the SomaLogic team for their support and the Meso Scale Discovery team for generous support with seroconversion assays. We are grateful to Michele Edelmann and the Health Data Compass team for the clinical data.

## AUTHOR CONTRIBUTIONS

All authors were involved in experimental design, data collection and/or analysis, and writing and/or review of the manuscript.

## SUPPLEMENTARY FIGURES

**Figure S1, related to Figure 1.**
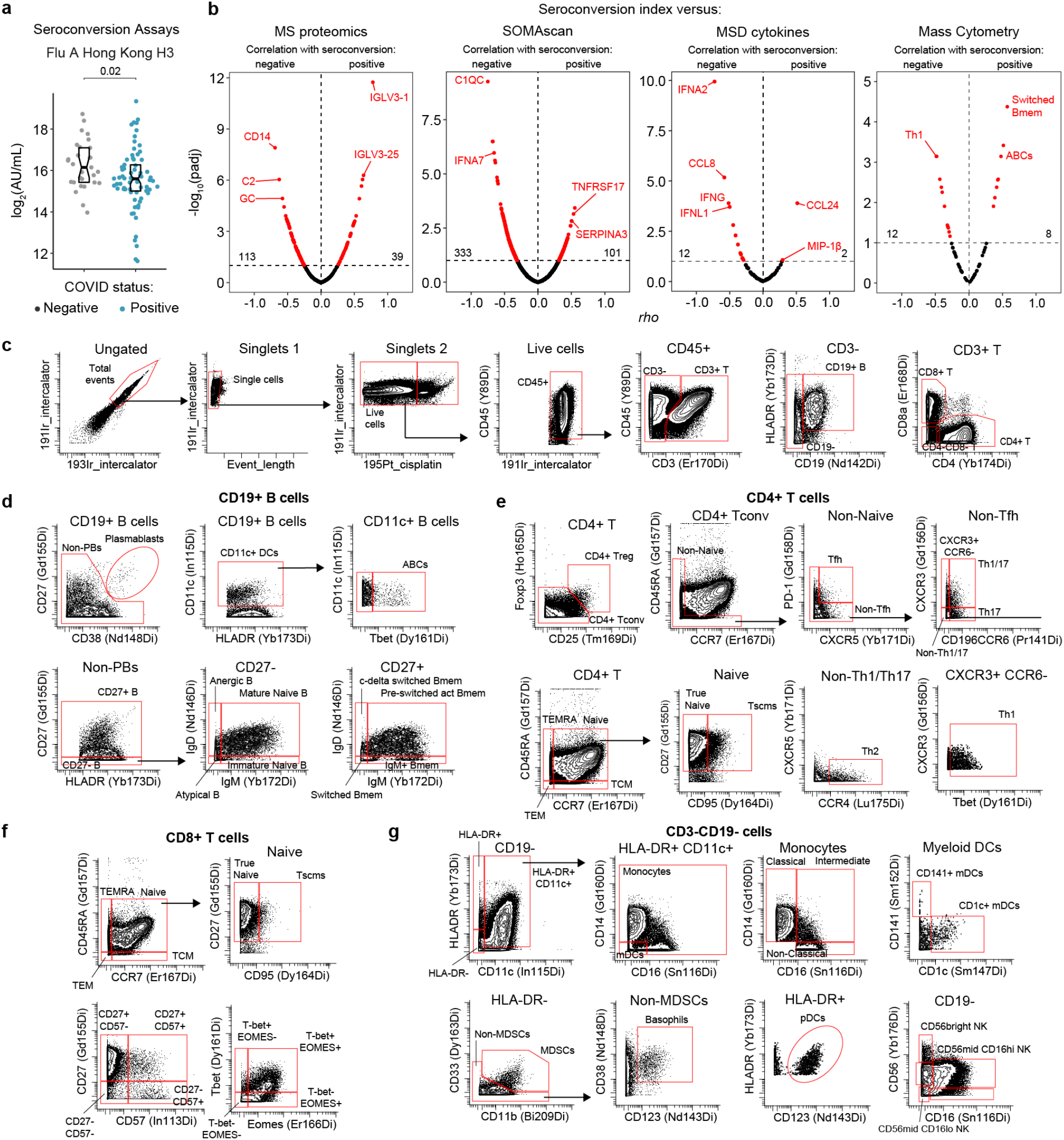
Biosignatures of seroconversion among hospitalized COVID19 patients. **a**. Meso Scale Discovery (MSD) assays show no elevation in levels of circulating antibodies against the Flu A Hong Kong H3 virus strain among COVID19 patients. Data are presented as modified Sina plots with boxes indicating median and interquartile range. Number above brackets is p-value for Mann-Whitney test. **b**. Volcano plots for Spearman correlations between seroconversion indices and circulating plasma proteins detected by mass spectrometry (MS), SOMAscan^®^ assays, MSD assays, as well as immune cell subsets detected by mass cytometry (MC) among all live cells. X axes show Spearman *rho* values. Y axes show -log_10_ p-values adjusted with Benjamini-Hochberg method. Dashed vertical line indicates *rho* = 0. Dashed horizontal line indicates the statistical cut off of false discovery rate (FDR)=10 (q=0.1). **c-g**. Representation of gating strategy employed during mass cytometry analysis of peripheral immune cell lineages. In (c), single live cells were gated for CD45+ staining followed by gating into T cells (CD3+), B cells (CD3-CD19+), and CD4+ and CD8+ T cells subsets. In (d), B cells were further gated into the indicated subsets. In (e) and (f), CD4+ and CD8+ T cell lineages were further characterized in the indicated subsets. Panel (g) shows gating for the indicated myeloid subsets and Natural Killer (NK) cells.

**Figure S2, related to Figure 2.**
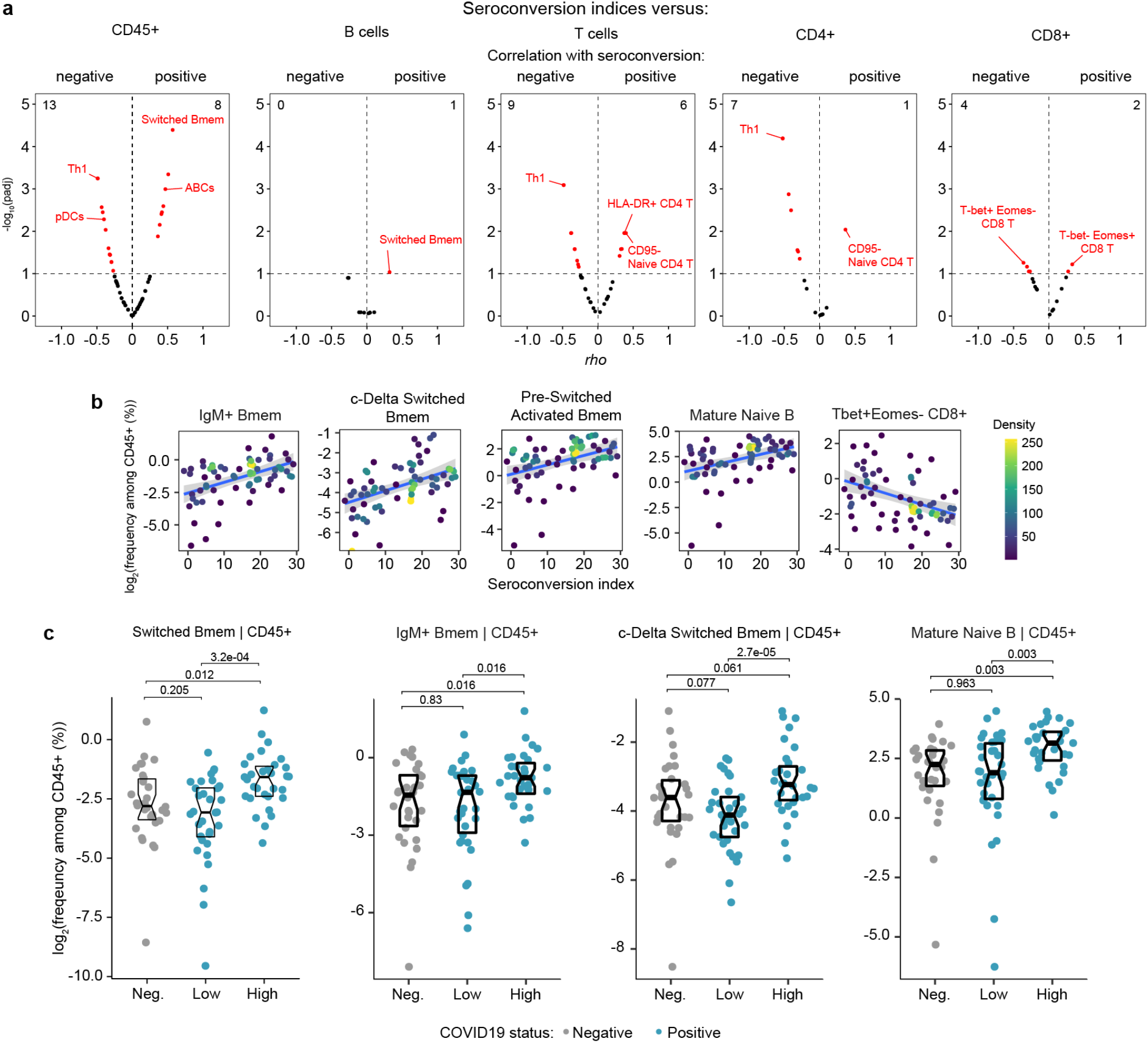
Immune cell signatures of seroconversion. **a**. Volcano plots displaying the correlations between seroconversion indices and circulating levels of immune cell subsets detected by mass cytometry (MC). X axes show Spearman *rho* values. Y axes show -log_10_ p-values adjusted with Benjamini-Hochberg method. Dashed vertical line indicates *rho* = 0. Dashed horizontal line indicates a false discovery rate (FDR) threshold of 10% (Q=0.1). Correlations were calculated for immune cell subsets measured among all live CD45+ cells (far left), all B cells, all T cells, CD4+ T cells, and CD8+ T cells (far right). **b**. Scatter plots for indicated cell types against seroconversion indices. Values shown are derived from frequency among all CD45+ cells. Points are colored by density; lines represent linear model fit with 95% confidence interval. **c**. Sina plots for indicated immune cell types comparing controls (Negative, Neg.) to COVID19 patients divided into seroconversion low (Low) and high (High) status. Data are presented as modified Sina plots with boxes indicating median and interquartile range. Number above brackets is Q-value for Mann-Whitney tests.

**Figure S3, related to Figure 3.**
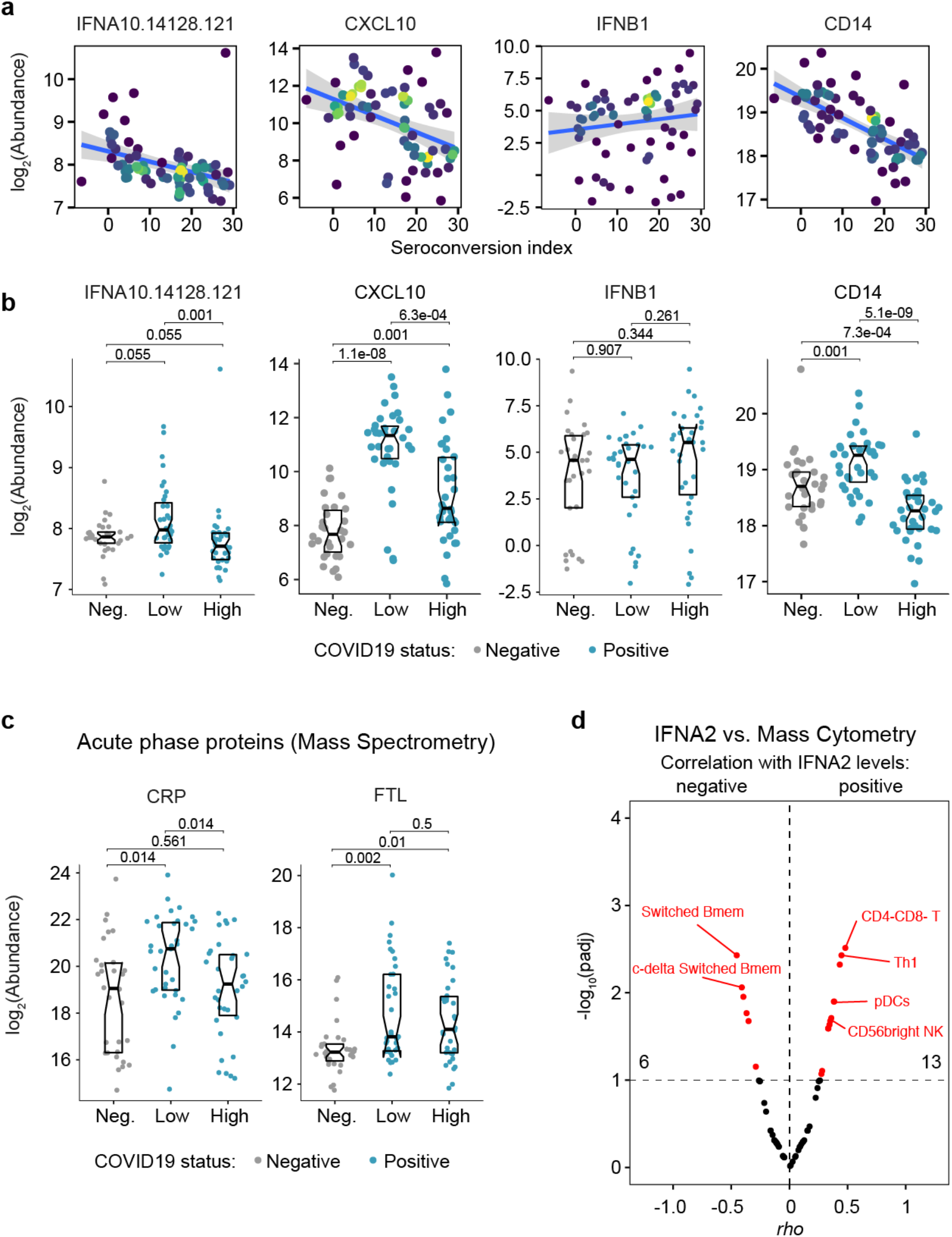
Seroconversion associates with differential abundance of circulating immune factors. **a-b**. XY scatter plots (a) and Sina plots (b) for select circulating immune factors. Points in (a) are colored by density; lines represent linear model fit with 95% confidence interval. Data in (b) are presented as modified Sina plots with boxes indicating median and interquartile range. Number above brackets is p-value for Mann-Whitney tests. **c**. Sina plots for the acute phase proteins CRP and FTL (Ferritin Light Chain) detected by MS comparing controls (Neg., negative) to COVID19 patients divided into seroconversion low (Low) and high (High) status. Data are presented as in b. **d**. Volcano plot showing associations between circulating levels of IFNA2 measured by MSD versus immune cell subsets among all live peripheral blood mononuclear cells. X axes show Spearman *rho* values. Y axes show -log_10_ p-values adjusted with Benjamini-Hochberg method. Dashed vertical line indicates *rho* = 0. Dashed horizontal line indicates a false discovery rate (FDR) threshold of 10% (Q=0.1).

**Figure S4, related to Figure 4.**
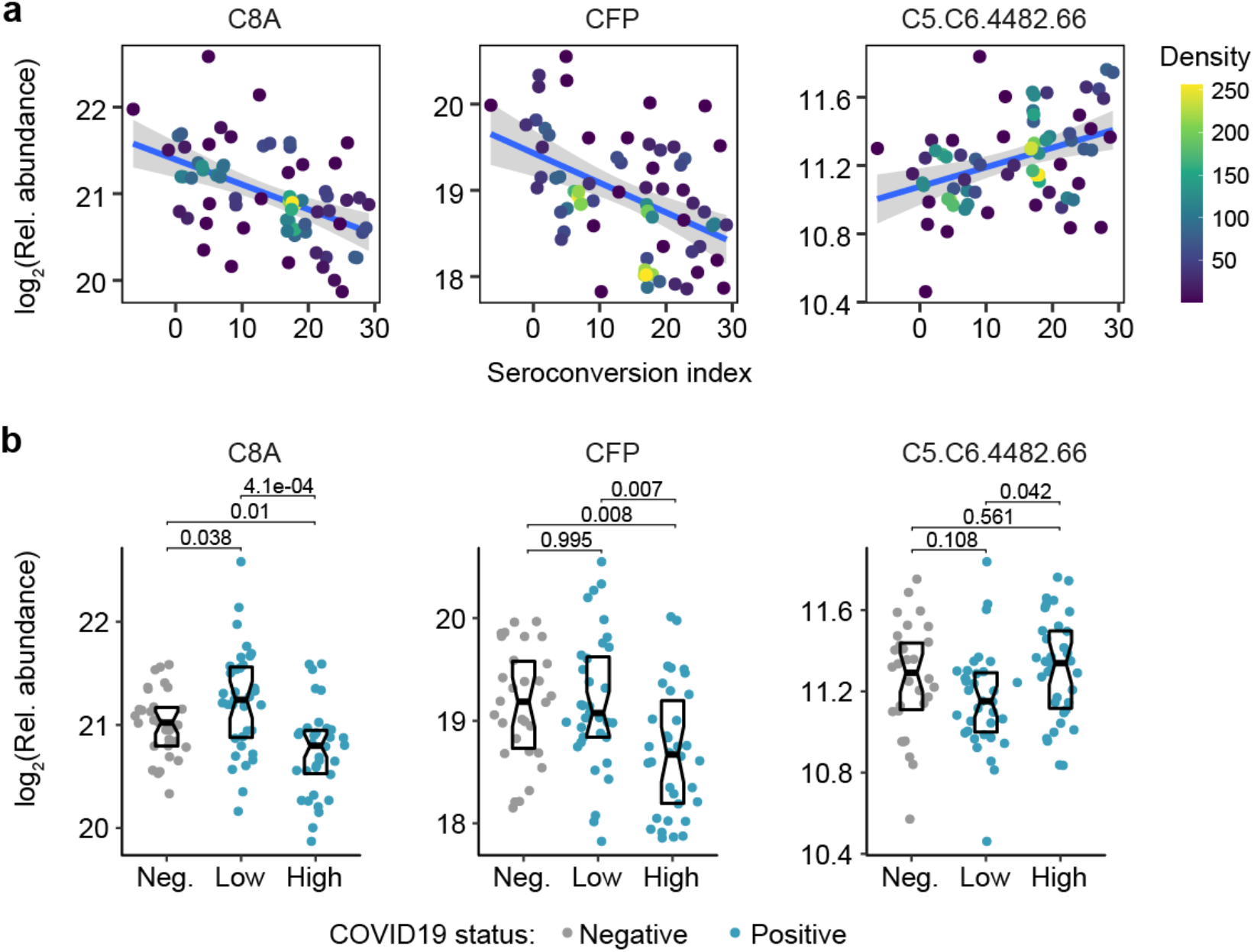
Seroconversion associates with decreased markers of systemic complement activation. **a-b**. XY scatter plots (a) and Sina plots (b) for select complement factors significantly associated, either positively or negatively, with seroconversion indices among COVID19 patients. Points in (a) are colored by density; lines represent linear model fit with 95% confidence interval. Data in (b) are presented as modified Sina plots with boxes indicating median and interquartile range comparing controls (Neg., negative) to COVID19 patients divided into seroconversion low (Low) and high (High) status. Number above brackets is p-value for Mann-Whitney tests.

**Figure S5, related to Figure 5.**
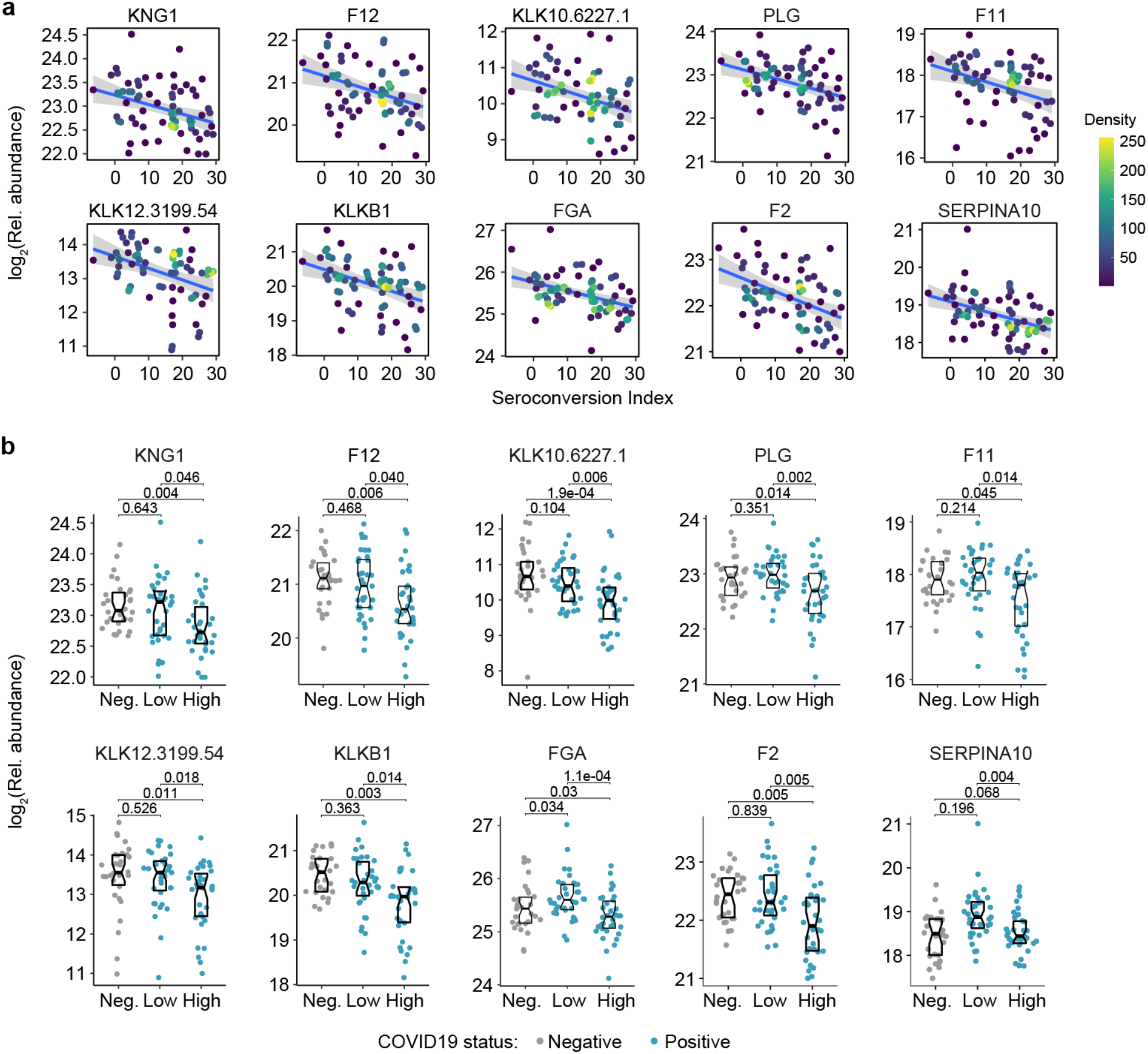
Seroconversion associates with remodeling of the hemostasis network. **a-b**. XY scatter plots (a) and Sina plots (b) for select factors involved in control of hemostasis significantly associated, either positively or negatively, with seroconversion indices among COVID19 patients. Points in (b) are colored by density; lines represent linear model fit with 95% confidence interval. Data in (b) are presented as modified Sina plots with boxes indicating median and interquartile range comparing controls (Neg., negative) to COVID19 patients divided into seroconversion low (Low) and high (High) status. Number above brackets is p-value for Mann-Whitney tests.

**Figure S6, related to Figure 6.**
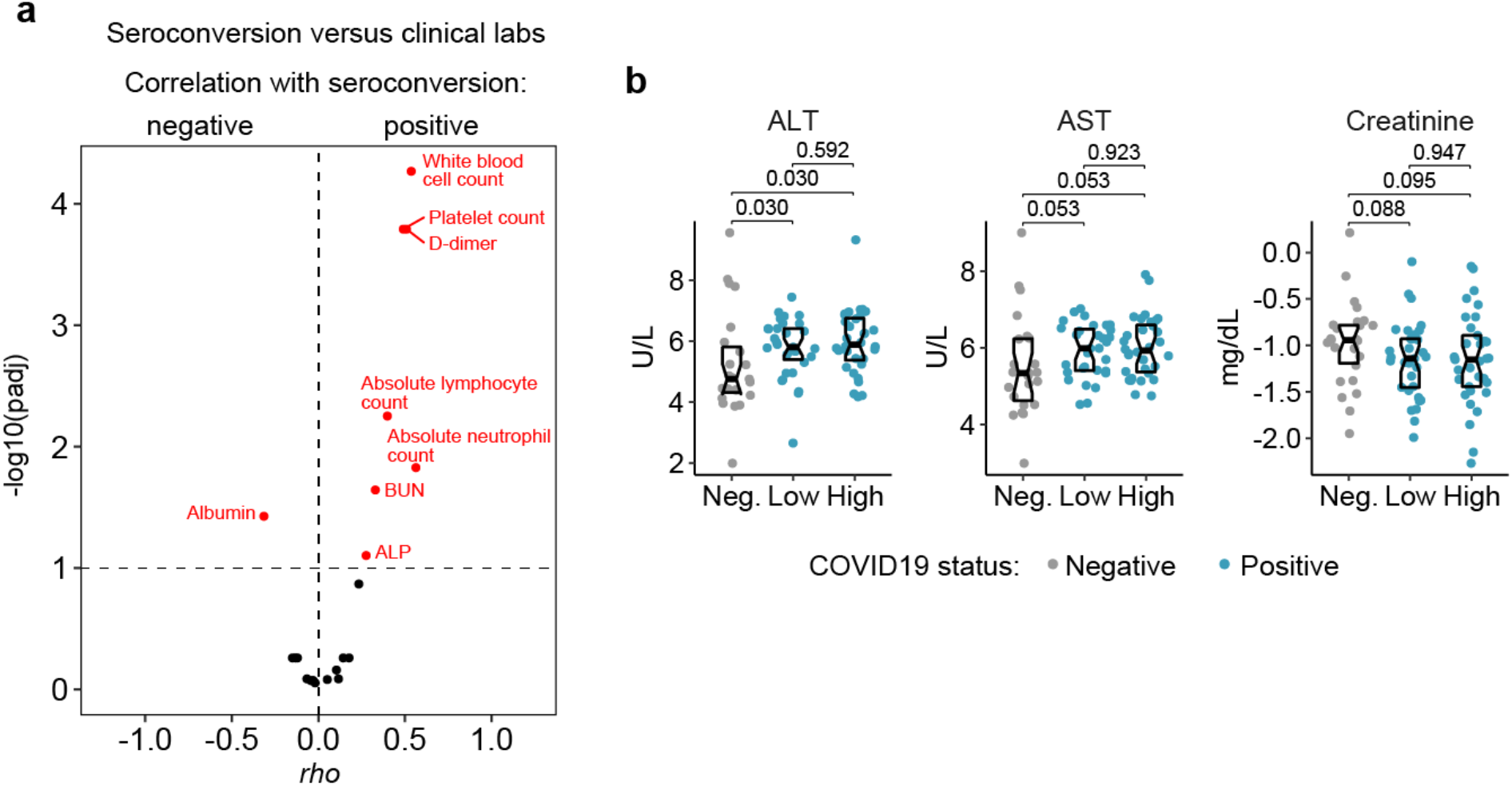
Seroconversion associates with recovery of blood cell counts and hypoalbuminemia. **a**. Volcano plot displaying the correlations between seroconversion indices and clinical laboratory values. X axes show Spearman *rho* values. Y axes show -log_10_ p-values adjusted with Benjamini-Hochberg method. Dashed vertical line indicates *rho* = 0. Dashed horizontal line indicates a false discovery rate (FDR) threshold of 10% (Q=0.1). **b**. Sina plots for select clinical laboratory values. Data in are presented as modified Sina plots with boxes indicating median and interquartile range comparing controls (Neg., negative) to COVID19 patients divided into seroconversion low (Low) and high (High) status. Number above brackets is p-value for Mann-Whitney tests.

## SUPPLEMENTARY FILES LEGENDS

**Supp. File S1. Cohort characteristics**. Table summarizing cohort characteristics. Information pertaining less than 10% of the cohort is indicated as <10% to prevent potential reidentification. **Supp. File S2. Seroconversion versus mass spectrometry proteomics**. Results of Spearman correlation analysis between seroconversion indices and proteins identified by Mass Spectrometry. Column A indicates the protein name, column B indicates the SwissProt ID, column C indicates the Spearman *rho* value, column D indicates the p value, and column E indicates the adjusted p value using the Benjamini-Hochberg method.

**Supp. File S3. Seroconversion versus SOMAcan**^**®**^ **proteomics**. Results of Spearman correlation analysis between seroconversion indices and proteins identified by SOMAscan^®^ technology. Column A indicates the SOMAmer identification number, column B indicates the target protein recognized by the SOMAmer, column C indicate the SwissProt ID, column D indicates the gene symsol, column E indicates the Spearman *rho* value, column F indicates the p value, and column G indicates the adjusted p value using the Benjamini-Hochberg method.

**Supp. File S4. Seroconversion versus MSD cytokine profiling**. Results of Spearman correlation analysis between seroconversion indices and cytokines, chemokines and other immune factors measured by multiplex immunoassays using Meso Scale Discovery (MSD) technology. Column A indicates the MSD analyte name, column B indicates the Spearman *rho* value, column C indicates the p value, and column D indicates the adjusted p value using the Benjamini-Hochberg method.

**Supp. File S5. Seroconversion versus mass cytometry**. Results of Spearman correlation analysis between seroconversion indices and immune cell subsets measured by mass cytometry (MC). Separate tabs are used for the results obtained from different parent lineages: all live cells, all CD45+ live cells, B cells, T cells, CD4+ T cells, CD8+ T cells, myeloid dendritic cells (mDCs) and monocytes. In each tab, column A indicates the population measured, column B indicates parent lineage, column C indicates the Spearman *rho* value, column D indicates the p value, and column E indicates the adjusted p value using the Benjamini-Hochberg method.

**Supp. File S6. Seroconversion versus immune factors**. Results of Spearman correlation analysis between seroconversion indices and circulating immune factors measured by mass spectrometry, SOMAscan® assays, or multiplex immunoassays with MSD technology. Column A indicates the analyte name in the platform, column B indicates the name used for display in the corresponding figure, column C indicates the platform used to measure the indicated analyte, column D indicates the Spearman *rho* value, column E indicates the p value, and column F indicates the adjusted p value using the Benjamini-Hochberg method.

**Supp. File S7. IFNA2 versus mass cytometry**. Results of Spearman correlation analysis between levels of IFNA2 measured by multiplex immunoassays using MSD technology versus immune cell subsets measured by mass cytometry. Column A indicates the immune cell population among all live cells, column B indicates the lineage used to calculate cell frequencies, column C indicates the Spearman *rho* value, column D indicates the p value, and column E indicates the adjusted p value using the Benjamini-Hochberg method.

**Supp. File S8. Seroconversion versus complement**. Results of Spearman correlation analysis between seroconversion indices and components of the complement pathways measured by mass spectrometry or SOMAscan^®^ assays. Column A indicates the unique identifier within the platform, column B indicates the name use for display in the corresponding figure, column C indicates the platform used to measure the indicated analyte, column D indicates the Spearman *rho* value, column E indicates the p value, and column E indicates the adjusted p value using the Benjamini-Hochberg method.

**Supp. File S9. Seroconversion versus hemostasis network**. Results of Spearman correlation analysis between seroconversion indices and factors involved in control of hemostasis measured by mass spectrometry or SOMAscan^®^ assays. Column A indicates the unique identifier within the platform, column B indicates the name use for display in the corresponding figure, column C indicates the platform used to measure the indicated analyte, column D indicates the Spearman *rho* value, column E indicates the p value, and column E indicates the adjusted p value using the Benjamini-Hochberg method.

**Supp. File S10. Seroconversion versus clinical laboratory values**. Results of Spearman correlation analysis between seroconversion indices and clinical laboratory values closest to the time of the research blood draw. Column A indicates the clinical laboratory parameter, column B indicates the Spearman *rho* value, column C indicates the p value, and column D indicates the adjusted p value using the Benjamini-Hochberg method.

**Supp. File S11. Mass cytometry antibody table**. List of antibodies used in mass cytometry. Column A indicates the antibody target, column B indicates the element conjugated to the antibody, column C indicates the mass of the element, column D indicates the manufacturer, column E indicates the catalog number, column F indicates the clone number, and column G indicates the type of stain protocol used (fixed, live or fixed with permeabilization).

**Supp. File S12. Immune cell type definition**. List of immune cell subsets defined by mass cytometry. Column A indicates the population identified, column B indicates definition based on gating strategy employed, and column C indicates the parent lineage.

